# Immunologic predictors of vaccine responsiveness in patients with lymphoma and CLL

**DOI:** 10.1101/2023.09.26.23295903

**Authors:** Elise A. Chong, Kingsley Gideon Kumashie, Emeline R. Chong, Joseph Fabrizio, Aditi Gupta, Jakub Svoboda, Stefan K. Barta, Kristy M. Walsh, Ellen B. Napier, Rachel K. Lundberg, Sunita D. Nasta, James N. Gerson, Daniel J. Landsburg, Joyce Gonzalez, Andrew Gaano, Madison E. Weirick, Christopher M. McAllister, Moses Awofolaju, Gavin N. John, Shane C. Kammerman, Josef Novaceck, Raymone Pajarillo, Kendall A. Lundgreen, Nicole Tanenbaum, Sigrid Gouma, Elizabeth M. Drapeau, Sharon Adamski, Kurt D’Andrea, Ajinkya Pattekar, Amanda Hicks, Scott Korte, Harsh Sharma, Sarah Herring, Justine C. Williams, Jacob T. Hamilton, Paul Bates, Scott E. Hensley, Eline T. Luning Prak, Allison R. Greenplate, E. John Wherry, Stephen J. Schuster, Marco Ruella, Laura A. Vella

## Abstract

Patients with B-cell lymphomas have altered cellular components of vaccine responses due to the malignancies and therapies. The optimal timing of vaccine administration relative to chemotherapy and immunotherapy remains unknown. The SARS-CoV-2 vaccine campaign created a unique opportunity to gather insights into vaccine timing because patients were challenged with a novel antigen across multiple phases of lymphoma management. We studied retrospective and prospective cohorts of patients with lymphoma and CLL who received an mRNA-based vaccine and paired serologic response with treatment dates, clinical immune parameters, and deep immunophenotyping. Reduced serologic response was observed more frequently during active therapies but nonresponders were also identified within observation and post-treatment groups. Clinical immunologic profiling demonstrated that total IgA and IgM near the time of vaccination correlated with ability to coordinate vaccine response. In individuals treated with CART-19, high-parameter immunophenotyping demonstrated that nonresponse was associated with reduced participation in B cell clusters and clusters of T follicular helper cells required for vaccine response. Together these data suggest that predictors of vaccine responsiveness vary by disease and therapeutic group. Further studies of immune health during and after cancer therapies will allow clinicians to individualize the timing of vaccines and define immunologic vulnerabilities.

## INTRODUCTION

The development of antibody responses after vaccination is considered a hallmark of immunogenicity for most vaccines^1,2^. In lymphoma and CLL, vaccine immunogenicity is particularly at risk, because both the malignancies and therapies directly impact B cells. In particular, B-cell-directed therapies such as monoclonal antibodies, Bruton tyrosine kinase (BTK) inhibitors, and anti-CD19 chimeric antigen receptor T cells (CART-19) drastically reduce B-cell numbers and function ^3–6^. There is currently no test to determine whether the immune system of a patient who is untreated, undergoing therapy or recovering immune system after therapy for lymphoma and CLL is able to coordinate a vaccine response.

Prior to 2020, studies of vaccine responsiveness during lymphoma therapy typically focused on influenza, *Streptococcus pneumoniae* and varicella zoster vaccine responses ^7–10^. These data are critical to understanding immune health of patients with lymphoma, but they primarily studied a memory response in the context of circulating vaccine-specific antibodies and potentially antigen-specific memory B cells. The response to a novel antigen is different and typically requires germinal center formation ^11,12^. Distinguishing new versus memory responses after a boost has been challenging ^13^, and therefore germinal-center dependent responses to novel antigens had been essentially unstudied in the setting of cancer therapies. The SARS-CoV-2 vaccine campaign therefore provided a singular opportunity to ask whether individuals across multiple types of lymphoma and CLL and across different stages of therapy would be able to coordinate a *de novo* antibody response against a novel antigen.

Studies of SARS-CoV-2 vaccine response in lymphoma and CLL have demonstrated heterogeneous vaccine immunogenicity, with variations observed depending on therapeutic modality and lymphoma subtype ^14–23^. Indeed, individuals with CLL and non-hodgkin lymphoma (NHL) treated with B-cell targeting therapies appeared to have particularly low serologic vaccine responses within 12 months of vaccination; the UK PROSECO study further highlighted the disconnect between serologic response and T cell responses, with T cell responses increasing over time despite lack of serologic response ^18,24–26^. Still, despite the therapeutic and disease-related barriers to coordinating B:T cell cross-talk required for serologic response, many individuals with lymphoma and CLL successfully developed IgG specific to this novel antigen.

The heterogeneity in SARS-CoV-2 vaccine response in lymphoma and CLL provided an opportunity to ask which immunological features predict whether the suppressed or emerging immune system is ready to coordinate a response to novel antigen. Further, the breadth of immunized individuals allowed for an assessment of predictors of vaccine responsiveness across multiple B cell-altering contexts. We used high-resolution clinical metadata to ask whether treatment type, time from treatment, or clinical measures of immunologic health correlated with the ability to coordinate a serologic vaccine response. We then performed cytometry by time of flight (CyTOF) on whole blood to determine whether higher dimensional measures of immune cell subsets could predict vaccine readiness. Our results indicate that individuals with lymphoma and CLL had varying responses to vaccination with reduced or absent serologic responses occurring even in the absence of active treatment and in patients who were treatment naive. Recovery of vaccine responsiveness correlated with time after receipt of B cell directed antibody therapies, but clinical and experimental measures of immune health varied by group.

## METHODS

### Patients and healthy donors

The study included 3 cohorts immunized with Pfizer-BioNTech or Moderna mRNA vaccines: (1) a prospective cohort of patients with NHL and CLL (research cohort); (2) a prospective cohort of healthy adults collected as previously published ^27,28^ with samples undergoing identical processing and antibody measurement alongside the research cohort (healthy cohort); (3) a retrospective observational cohort of patients with NHL and CLL (clinical cohort). Additional cohort details are in **Supplementary Methods**.

### Clinical Data Abstraction

Patient level clinical data were abstracted from the electronic medical record by the investigators as outlined in **Supplementary Methods**

### Research and Healthy Cohort Blood Collection and Serology

Peripheral blood was collected 0-90 days prior to first vaccination, 1-3 weeks after vaccine dose 1, 1-5 months after vaccine dose 2, and, when applicable, 1-2 weeks after third immunization. Enzyme-Linked Immunosorbent Assay (ELISA) was performed on plasma obtained from blood samples, as described ^29,30^ and details are in Supplementary Methods.

### Clinical Cohort Blood Collection and Serology

Peripheral blood was collected for serologic testing at the timing and discretion of the treating physician and details of sample processing and analyses are in **Supplementary Methods**. Results were reported in arbitrary units (AU). Reference ranges were established using samples from patients with COVID-19 and pre-2019 controls. The reference ranges were: negative (≤0.3), equivocal (0.301-0.699), positive (≥0.7).

### Whole Blood CyTOF

Whole blood (300µl) sampled between 14 days and 4 months after the second dose of vaccination was added to the Maxpar Direct Immune Profiling Assay kit (Standard BioTools). Details in **Supplementary Methods.**

### High dimensional CyTOF data analysis

Data was analyzed using OMIQ cytometry analysis platform (https://app.omiq.ai/). Details are in the **Supplementary Methods**.

### Statistical analyses

See **supplementary Methods**.

## RESULTS

### Patients with lymphoma have impaired vaccine responses

To determine how immune responses in patients with lymphomas and CLL compared to those of healthy adults, we enrolled 71 patients with NHL/CLL who planned to receive or were within 1 month of receipt of SARS-CoV-2 immunization (**Table 1**). Control subjects were healthy adults who had blood collected, processed, and analyzed in parallel, and have been previously described^27^. Patients with NHL/CLL underwent sample collection at time points aligned with clinical visits (**Figure 1A**); 24 and 65 patients had specimens available after the first and second vaccine dose, respectively. Both FDA-authorized mRNA vaccines were included; 40 (56%) received Pfizer-BioNTech and 31 (44%) received Moderna (**Table 1**). At a median of 2.1 weeks after vaccine dose 1, patients with NHL/CLL demonstrated a reduced frequency of seroconversion (6/24 [25%] versus 32/33 [97%]) compared to healthy controls (**Figure 1B**, left panel). Six weeks (range: 0.5-23) after vaccine dose 2, the proportion of patients with lymphoma/CLL who developed detectable RBD-specific IgG antibodies increased, but remained significantly lower than in healthy controls (33/65 [51%] versus 32/32 [100%]) (**Figure 1B**, right panel). As a group, no B-cell lymphoma subtype was associated with a normal serologic vaccine response, compared to healthy controls, although some patients with NHL/CLL demonstrated normal or near-normal anti-RBD concentrations (**Figure 1C**).

**Figure 1:**
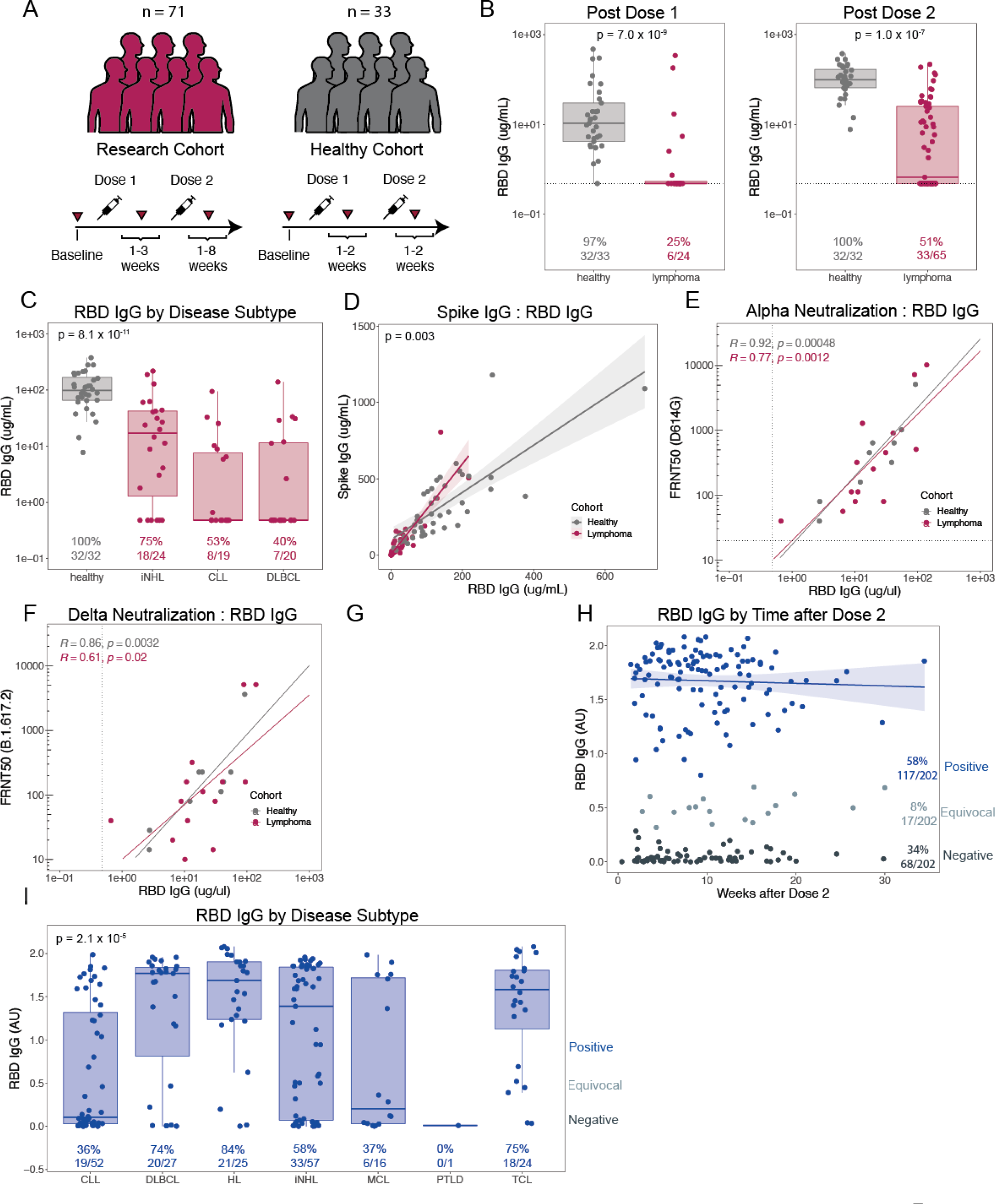
Serologic Response to Vaccination in Patients with Lymphoma/CLL. (**A**) Schema of research and healthy cohort blood draw timepoints. (**B**) Anti-RBD IgG after vaccine dose 1 and dose 2 in research and healthy cohorts. Dotted line reflects the limit of anti-RBD IgG detection. Proportion positive indicated at bottom. P-values derived from Fisher’s exact tests. (**C**) Anti-RBD IgG by disease subtype after second vaccination. P-value derived from Kruskal Wallis test of difference across all groups. The iNHL group was comprised of 17 patients not receiving active treatment and 7 active treatment patients, the CLL group comprised 8 patients receiving active treatment and 11 untreated patients, and the DLBCL group predominantly comprised patients status post CART-19 cells (N=16) while the other 4 patients were not receiving active treatment. (**D**) Correlation between anti-spike IgG and anti-RBD IgG in each cohort. P-value represents a test for difference between the two correlations using a linear model. (**E-F**) Correlation between RBD-IgG and pseudovirus neutralization titers for alpha variant (**E**, D614G) and delta variant (**F**, B.617.2), described as focus reduction neutralization titer 50% (FRNT50) in a subset of patients with detectable RBD-IgG. (**G**) Description of clinical cohort blood draw timepoints. (**H**) Cross-sectional distribution of anti-RBD IgG measured in the clinical assay by time of blood draw. Each individual is represented once. Dashed lines represent thresholds between positive samples, equivocal samples, and negative samples. The proportion in each category is at right. Trend line is for positive samples only. (**I**) Anti-RBD IgG by disease subtype in the clinical cohort. Proportion of positive patients indicated at bottom. P-value derived from Kruskal Wallis test of difference across all groups. *A.U. = arbitrary units; RBD= receptor-binding domain; CLL = chronic lymphoid leukemia; iNHL = indolent non-Hodgkin lymphoma; DLBCL = diffuse large B-cell lymphoma; HL = Hodgkin lymphoma; MCL = mantle cell lymphoma; TCL = T-cell lymphoma; PTLD = post-transplant lymphoproliferative disease*.

**Table 1.**
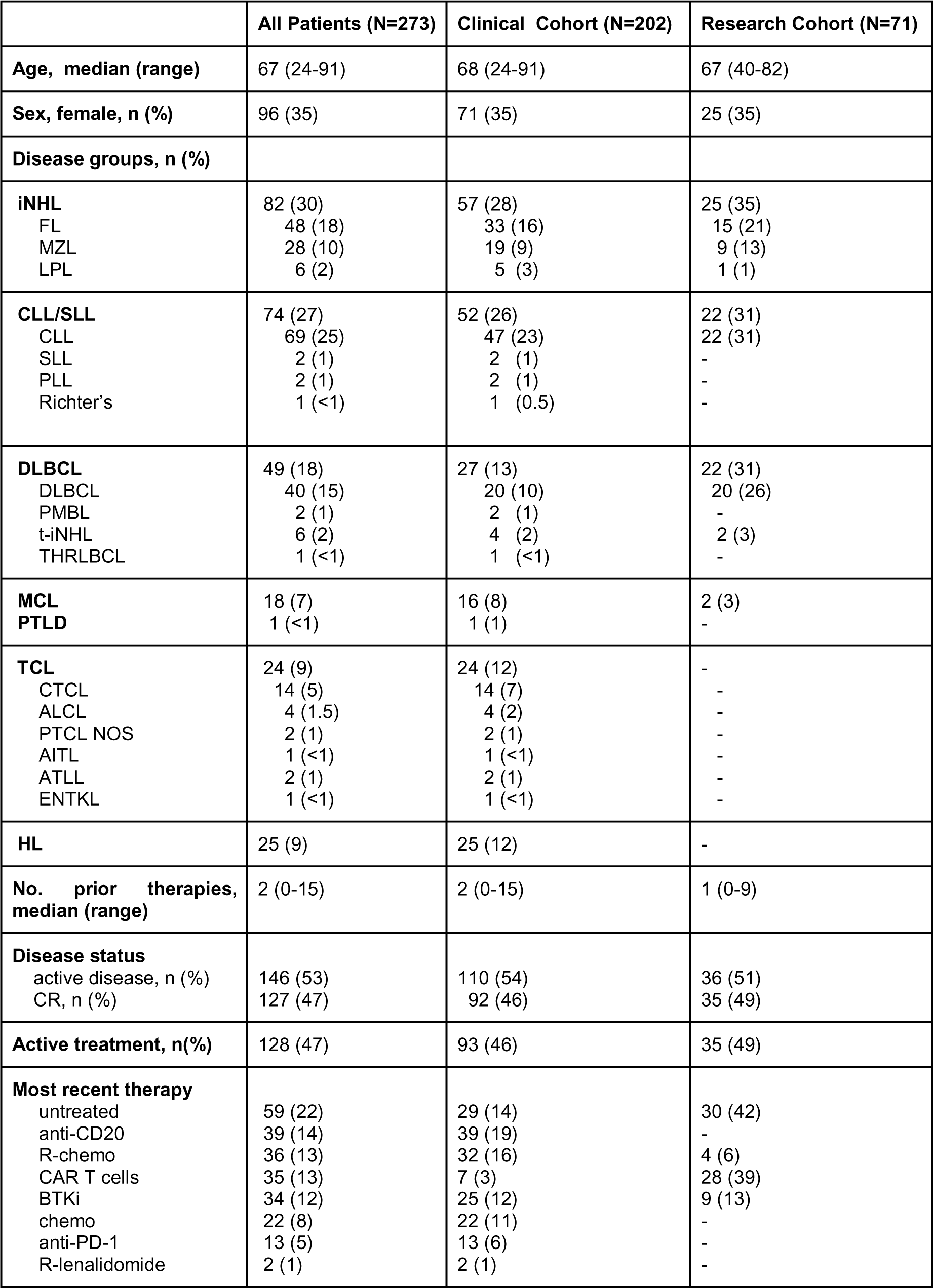
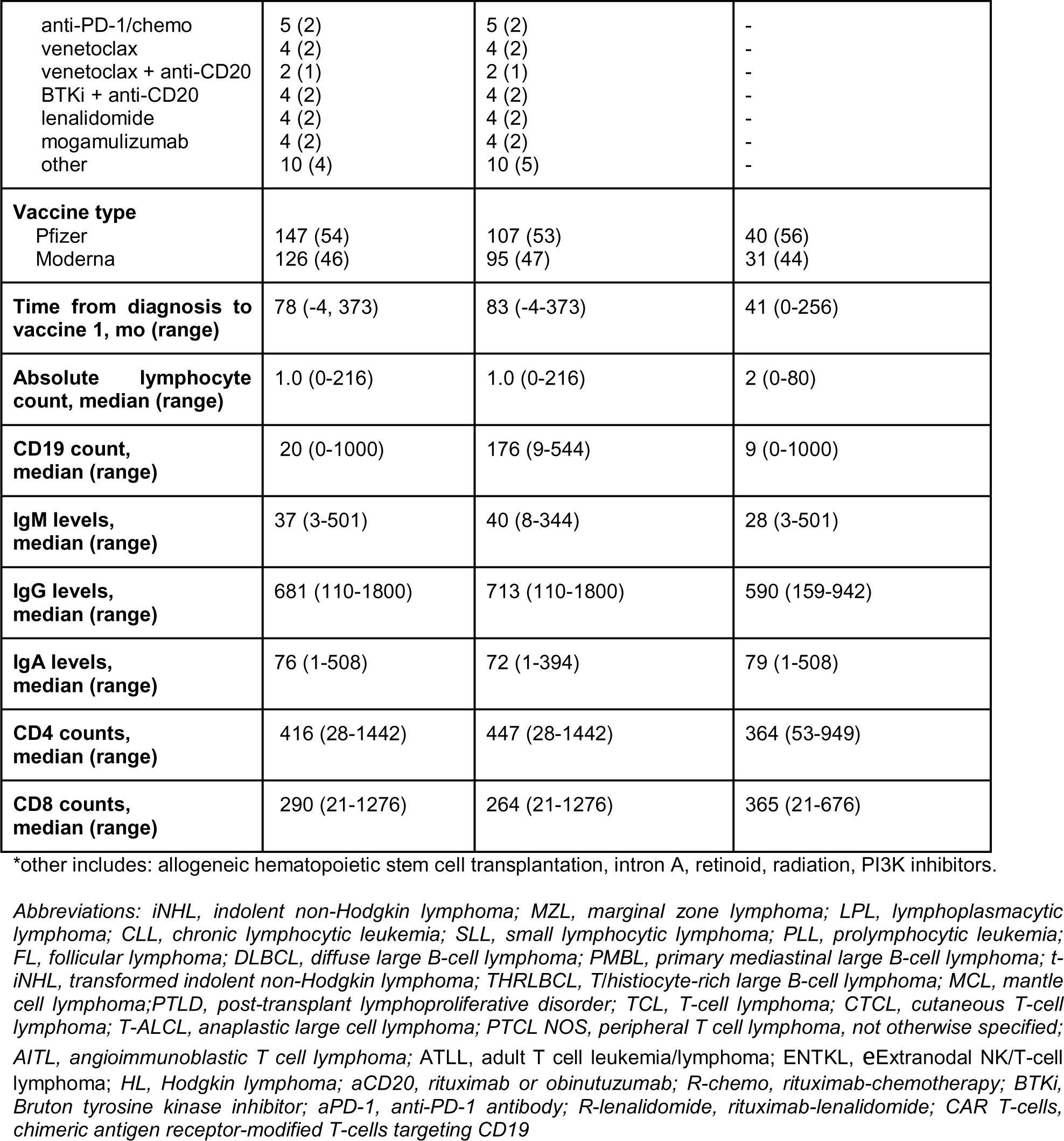
Patients’ Characteristics.

|  | All Patients (N=273) | Clinical Cohort (N=202) | Research Cohort (N=71) |
| --- | --- | --- | --- |
| **Age, median (range)** | 67 (24-91) | 68 (24-91) | 67 (40-82) |
| **Sex, female, n (%)** | 96 (35) | 71 (35) | 25 (35) |
| **Disease groups, n (%)** |  |  |  |
| **iNHL** | 82 (30) | 57 (28) | 25 (35) |
| FL | 48 (18) | 33 (16) | 15 (21) |
| MZL | 28 (10) | 19 (9) | 9 (13) |
| LPL | 6 (2) | 5 (3) | 1 (1) |
| **CLL/SLL** | 74 (27) | 52 (26) | 22 (31) |
| CLL | 69 (25) | 47 (23) | 22 (31) |
| SLL | 2 (1) | 2 (1) | - |
| PLL | 2 (1) | 2 (1) | - |
| Richter's | 1 (<1) | 1 (0.5) | - |
| **DLBCL** | 49 (18) | 27 (13) | 22 (31) |
| DLBCL | 40 (15) | 20 (10) | 20 (26) |
| PMBL | 2 (1) | 2 (1) | - |
| t-iNHL | 6 (2) | 4 (2) | 2 (3) |
| THRLBCL | 1 (<1) | 1 (<1) | - |
| **MCL** | 18 (7) | 16 (8) | 2 (3) |
| **PTLD** | 1 (<1) | 1 (1) | - |
| **TCL** | 24 (9) | 24 (12) | - |
| CTCL | 14 (5) | 14 (7) | - |
| ALCL | 4 (1.5) | 4 (2) | - |
| PTCL NOS | 2 (1) | 2 (1) | - |
| AITL | 1 (<1) | 1 (<1) | - |
| ATLL | 2 (1) | 2 (1) | - |
| ENTKL | 1 (<1) | 1 (<1) | - |
| **HL** | 25 (9) | 25 (12) | - |
| **No. prior therapies, median (range)** | 2 (0-15) | 2 (0-15) | 1 (0-9) |
| **Disease status** |  |  |  |
| active disease, n (%) | 146 (53) | 110 (54) | 36 (51) |
| CR, n (%) | 127 (47) | 92 (46) | 35 (49) |
| **Active treatment, n(%)** | 128 (47) | 93 (46) | 35 (49) |
| **Most recent therapy** |  |  |  |
| untreated | 59 (22) | 29 (14) | 30 (42) |
| anti-CD20 | 39 (14) | 39 (19) | - |
| R-chemo | 36 (13) | 32 (16) | 4 (6) |
| CAR T cells | 35 (13) | 7 (3) | 28 (39) |
| BTKi | 34 (12) | 25 (12) | 9 (13) |
| chemo | 22 (8) | 22 (11) | - |
| anti-PD-1 | 13 (5) | 13 (6) | - |
| R-lenalidomide | 2 (1) | 2 (1) | - |
| anti-PD-1/chemo | 5 (2) | 5 (2) | - |
| venetoclax | 4 (2) | 4 (2) | - |
| venetoclax + anti-CD20 | 2 (1) | 2 (1) | - |
| BTKi + anti-CD20 | 4 (2) | 4 (2) | - |
| lenalidomide | 4 (2) | 4 (2) | - |
| mogamulizumab | 4 (2) | 4 (2) | - |
| other | 10 (4) | 10 (5) | - |
| **Vaccine type** |  |  |  |
| Pfizer | 147 (54) | 107 (53) | 40 (56) |
| Moderna | 126 (46) | 95 (47) | 31 (44) |
| **Time from diagnosis to vaccine 1, mo (range)** | 78 (-4, 373) | 83 (-4-373) | 41 (0-256) |
| **Absolute lymphocyte count, median (range)** | 1.0 (0-216) | 1.0 (0-216) | 2 (0-80) |
| **CD19 count, median (range)** | 20 (0-1000) | 176 (9-544) | 9 (0-1000) |
| **IgM levels, median (range)** | 37 (3-501) | 40 (8-344) | 28 (3-501) |
| **IgG levels, median (range)** | 681 (110-1800) | 713 (110-1800) | 590 (159-942) |
| **IgA levels, median (range)** | 76 (1-508) | 72 (1-394) | 79 (1-508) |
| **CD4 counts, median (range)** | 416 (28-1442) | 447 (28-1442) | 364 (53-949) |
| **CD8 counts, median (range)** | 290 (21-1276) | 264 (21-1276) | 365 (21-676) |

\*other includes: allogeneic hematopoietic stem cell transplantation, intron A, retinoid, radiation, PI3K inhibitors.
Abbreviations: iNHL, indolent non-Hodgkin lymphoma; MZL, marginal zone lymphoma; LPL, lymphoplasmacytic lymphoma; CLL, chronic lymphocytic leukemia; SLL, small lymphocytic lymphoma; PLL, prolymphocytic leukemia; FL, follicular lymphoma; DLBCL, diffuse large B-cell lymphoma; PMBL, primary mediastinal large B-cell lymphoma; t-iNHL, transformed indolent non-Hodgkin lymphoma; THRLBCL, T/histiocyte-rich large B-cell lymphoma; MCL, mantle cell lymphoma; PTLD, post-transplant lymphoproliferative disorder; TCL, T-cell lymphoma; CTCL, cutaneous T-cell lymphoma; T-ALCL, anaplastic large cell lymphoma; PTCL NOS, peripheral T cell lymphoma, not otherwise specified; AITL, angioimmunoblastic T cell lymphoma; ATLL, adult T cell leukemia/lymphoma; ENTCL, eExtranodal NK/T-cell lymphoma; HL, Hodgkin lymphoma; aCD20, rituximab or obinutuzumab; R-chemo, rituximab-chemotherapy; BTKi, Bruton tyrosine kinase inhibitor; aPD-1, anti-PD-1 antibody; R-lenalidomide, rituximab-lenalidomide; CAR T-cells, chimeric antigen receptor-modified T-cells targeting CD19

Although both anti-spike and anti-RBD antibodies correlate with neutralizing antibody levels, most neutralizing antibodies target a portion of RBD^32^, and the ratio of anti-RBD to total anti-spike IgG can roughly estimate the quality of the antibody response. In lymphoma patients who generated an antibody response, the anti-RBD to anti-spike ratio was significantly lower than in healthy controls (**Figure 1D**). However, it remained unclear whether functional differences exist among patients with RBD-binding IgG. Thus, we assessed neutralizing antibodies in a subset of patients with detectable RBD-specific IgG (**Supplemental Table 2**). For the alpha variant (D614G) of SARS-CoV-2, when RBD-specific IgG was detectable, serological responses correlated strongly with neutralizing antibody titers (focus reduction neutralization titer 50% or FRNT50), and correlations were similar across healthy and lymphoma cohorts (**Figure 1E**). When tested against the delta variant (B.1.617.2), the correlations were reduced in general, with some individuals having a discordant RBD-IgG and FRNT50 status. The correlations were less similar between cohorts but were not statistically different (**Figure 1F**). Together, these data suggested that the rate of seroconversion and the concentration of RBD-specific IgG are decreased in lymphoma/CLL but, when present, RBD-specific IgG correlated with presence of neutralizing antibodies.

To understand differences between lymphoma/CLL subtypes more broadly, we examined the clinical cohort with lymphoma/CLL for whom anti-RBD IgG was measured using a clinical assay. Two hundred two patients were enrolled (**Table 1**, *Clinical cohort*). All received an mRNA-based SARS-CoV-2 vaccine. Patient samples were obtained at a median of 9 weeks (range: 0.5-34 weeks) after the second vaccine dose; 117/202 patients (58%) seroconverted (**Figure 1H**). In patients who developed an antibody response following vaccination, responses appeared durable based on cross-sectional analysis over the first 30 weeks (**Figure 1H**, blue line). As with the research cohort, anti-RBD IgG responses varied by lymphoma subtype (**Figure 1I**).

### Therapeutic class and time from therapy completion differentially impact serologic responses to SARS-CoV-2 vaccines

Decreased vaccine responses in patients with lymphoma and CLL could be due to the presence of malignancy and/or due to the effect of anti-cancer therapy. We, therefore, assessed the effect of active treatment on humoral response to SARS-CoV-2 vaccines (**Figure 2A**). As shown in **Table 1**, 93 of 202 patients (46%) of the clinical cohort were receiving active treatment (within 5 half-lives of therapeutic agent, 3 months of last chemotherapy dose, 6 months of last anti-CD20 antibody) prior to vaccination, or were status post CART-19 without progressive lymphoma. Active treatment was associated with a reduced frequency of seroconversion (34% versus 78%) (**Figure 2B**). Antibody responses were lower in patients receiving active treatment in CLL, indolent NHL, mantle cell lymphoma (MCL), and T-cell lymphoma (TCL), whereas there was no difference for Hodgkin Lymphoma (HL) or diffuse large B-cell lymphoma (DLBCL) patients (**Figure 2C**). For patients not receiving active treatment, individuals with CLL had significantly lower antibody levels after two doses of vaccination compared to those with DLBCL, iNHL, HL, and TCL.

**Figure 2:**
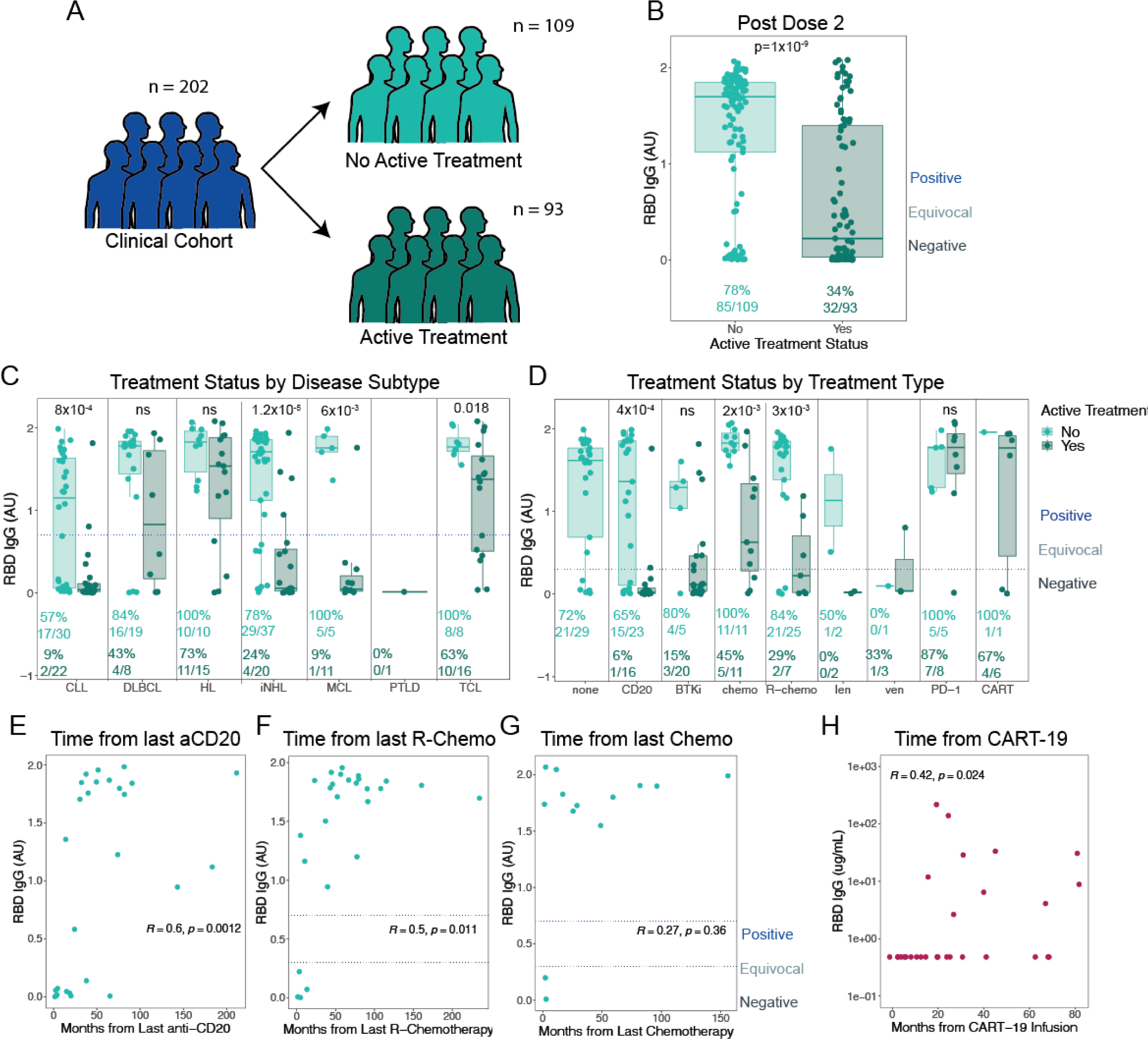
Active Treatment is Associated with Decreased Immunogenicity of SARS-CoV-2 vaccines. (**A**) Description of treated versus untreated patients within the clinical cohort. (**B**) Anti-RBD IgG after vaccine dose 2 in patients undergoing active treatment versus those not undergoing active treatment. P-value derived from Fisher’s exact test. Dashed lines represent thresholds between positive samples, equivocal samples, and negative samples, as indicated. (**C and D**) Anti-RBD IgG in active treatment versus no active treatment across disease subtypes (**C**) and across therapies (**D**). P-values indicated at the top, derived from Wilcoxon Rank Sum tests with corrections for multiple comparisons (**E-G**) Correlation between anti-RBD IgG in the clinical cohort (dark blue, A) by time from therapy with anti-CD20, R-chemotherapy, and chemotherapy. Horizontal dashed lines as in B. (**H**) Correlation between time from CART-19 infusion versus anti-RBD IgG in the research cohort. P-values in D-F are derived from Spearman correlation tests. *A.U. = arbitrary units; RBD= receptor-binding domain; CLL = chronic lymphoid leukemia; iNHL = indolent non-Hodgkin lymphoma, DLBCL = diffuse large cell B-cell lymphoma; HL = Hodgkin lymphoma; MCL = Mantle cell lymphoma. TCL = T-cell lymphoma; PTLD = post-transplant lymphoproliferative disease; CD20 = anti-CD20 monoclonal; BTKi = Bruton’s tyrosine kinase inhibitor; R-chemo = anti-CD20 with chemotherapy; len = lenalidomide; ven =venetoclax; PD-1 = therapy with anti-PD-1 monoclonal*

We next investigated the effect of specific therapies on seroconversion. **Table 1** describes most recently administered lymphoma therapies. Lack of response was observed in patients receiving anti-CD20 monoclonal antibodies, anti-CD20 based chemoimmunotherapy, and chemotherapy (**Figure 2D**). Notably, among patients receiving active treatment, we observed that patients receiving anti-PD1 antibody therapy had consistently high rates of vaccine response.

Monoclonal antibody therapy targeting CD20 and CART-19 are associated with a long half-life and prolonged B-cell depletion. We, therefore, assessed vaccine response as a function of time from the last anti-CD20 antibody dose and from CART-19 cells. Time from anti-CD20 therapy positively correlated with vaccine-induced anti-RBD IgG in both patients receiving anti-CD20 antibody therapy alone (**Figure 2E**) and concomitant anti-CD20-chemoimmunotherapy (**Figure 2F**). Conversely, time from chemotherapy alone was not correlated with vaccine response, suggesting that immune reconstitution was rapid and active treatment alone is the primary driver of non-response (**Figure 2G**).

Patients in remission after CART-19 therapy generally had poorer antibody responses for a significantly longer duration after completion of therapy. For the first 1-2 years after CART-19 infusion, anti-RBD IgG values were consistently negative. In some patients, undetectable vaccine responses occurred many years after CART-19 infusion, although other patients demonstrated seroconversion 1-2 years post-CART-19 infusion (**Figure 2H**). This observation is consistent with known timing of B cell recovery after CART-19 therapy^33^.

### Clinical measures of immune health and response to SARS-CoV-2 mRNA vaccination

We also hypothesized that patients with lymphoma/CLL might have differential responses to SARS-CoV-2 vaccination based on the baseline “health” of their immune system. Indeed, patients with normal concentrations of total immunoglobulins before vaccination were more likely to mount a detectable antibody response; however, this association was not evident for T cell or T cell subset counts (**Figures 3A-G**, top rows). Similarly, in analyses of correlation, absolute CD19 and immunoglobulin levels positively correlated RBD-specific IgG (**Figures 3A-G**, bottom rows). No measured immunologic parameter fully discriminated between response and non-response.

**Figure 3:**
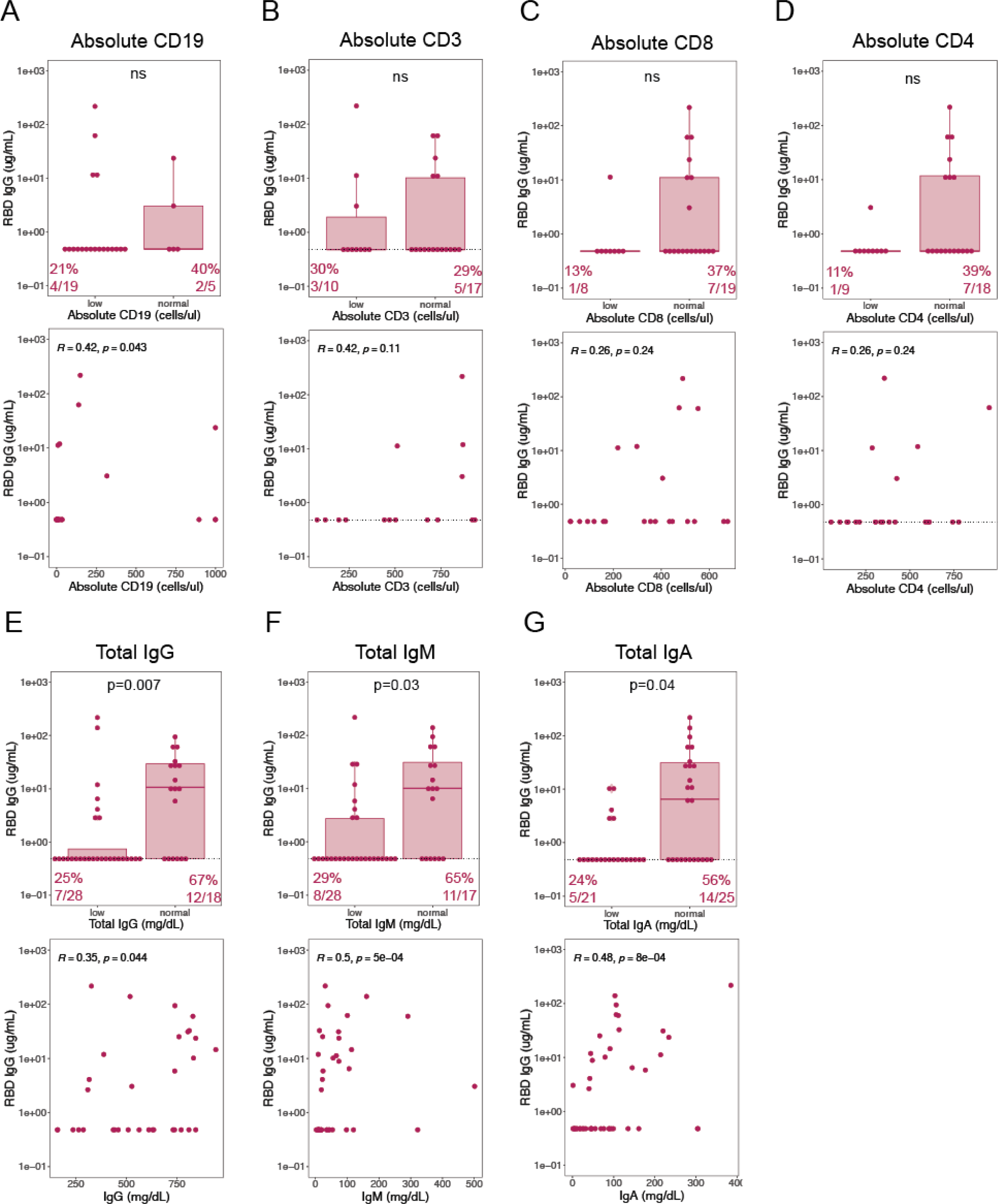
Baseline Immune Cell frequencies in the Blood were Associated with Vaccine Immunogenicity. (**A-G**, Top row) Research Cohort anti-RBD IgG by normal versus below normal clinical measures of immune status (top row, proportions of positive tests for normal and low clinical values noted at bottom. P-values are derived from Fisher’s exact tests. (**A-G**, bottom row) Correlation between anti-RBD IgG versus values of clinically measured immunologic parameters. P-values are derived from Spearman correlation tests.

### SARS-CoV-2 mRNA vaccine response in DLBCL patients post CAR T cell therapy is associated with B cell reconstitution

To gain a deeper understanding of immune predictive features in vaccine responders vs non-responders we performed a high-parameter immunologic assessment in individuals with DLBCL who were status post CART-19 infusion (DLBCL-CART-19). We assessed 14 individuals with DLBCL-CART-19 (5 responders and 9 non-responders, see **Supplemental Table 3**), and 23 healthy donor peripheral blood samples using CyTOF analysis. Data were analyzed using t-SNE, and FlowSOM approaches to define and quantify cell populations that differed between the responder and non-responders (**Figure 4A, B**). These approaches highlighted a loss of participating cells from cluster 3 in DLBCL-CART-19 vaccine non-responders when compared to healthy control and DLBCL-CART-19 responders (**Figures 4A-E**). This cluster expressed CD19 and CD20, representing B cells which accounted for 10-15% of B cells in responders compared to non-responders (**Figures 4C-E**). This B cell cluster expressed varying degrees of CD38, IgD, CCR6, CD45RA, HLA-DR and CD27, reflecting activated B cells (**Figures 4C-E**). Together these data are consistent with B-cells serving as a driving predictor of vaccine response in DLBCL-CART-19 patients.

**Figure 4:**
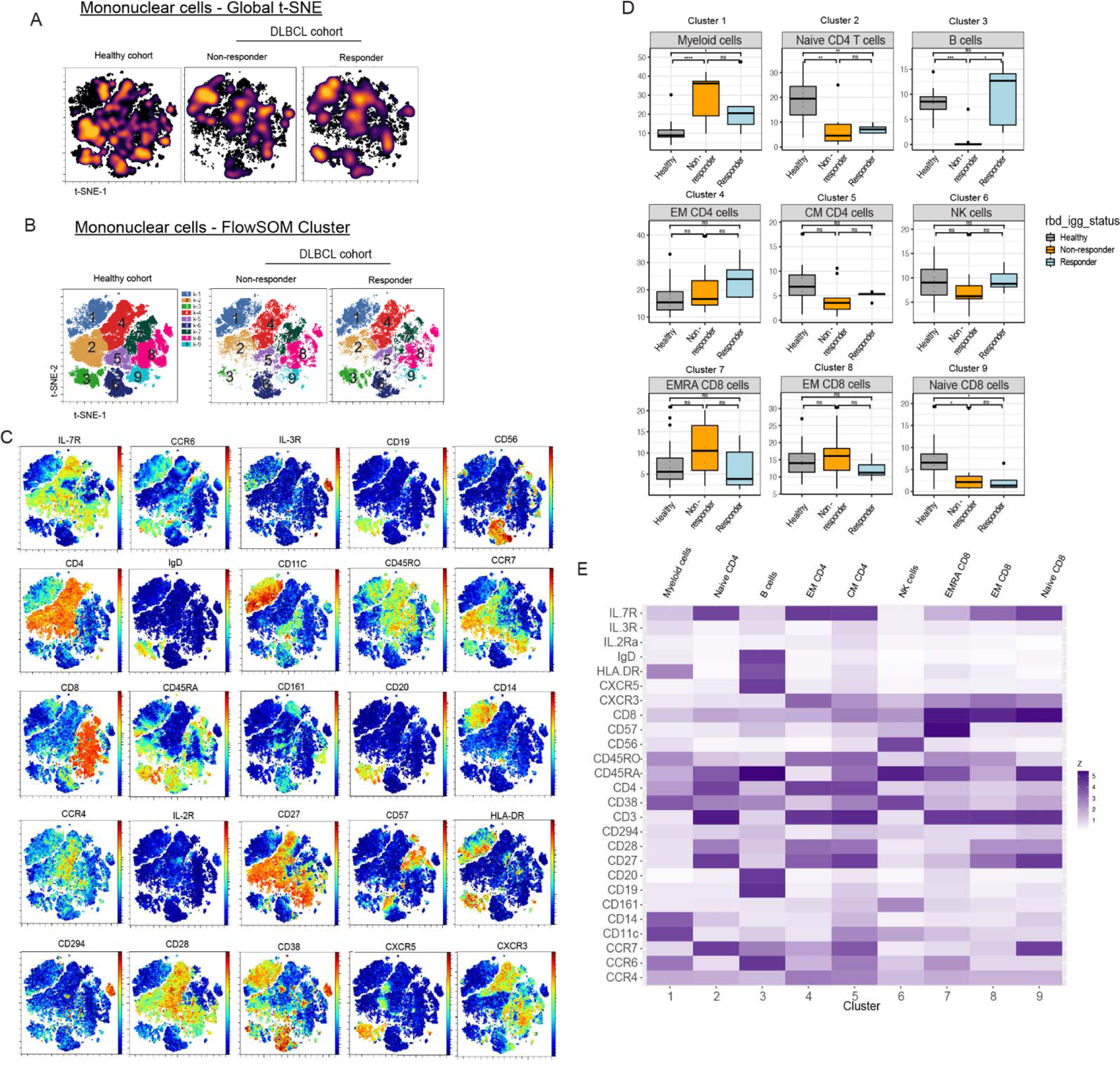
SARS-CoV-2 mRNA vaccine response in DLBCL post CAR T cell therapy is associated with B cell reconstitution. (**A**) Global t-SNE projection of mononuclear leukocytes for all pooled subjects from DLBCL responder and non-responders as well as healthy cohorts. (**B**) FlowSOM clusters of mononuclear cells (lymphocytes and monocytes) concatenated from each DLBCL cohort (non-responder(n=9) and responder (n=5)) and healthy cohort (n=23). (**C**) Projection of indicated protein onto t-SNE map. (**D**) Boxplot of mononuclear cell frequencies from each cohort in each FlowSOM cluster in (c). (**E**) Median fluorescence intensity (MFI) of each marker in each FlowSOM cluster (row scaled z-score). Significance was determined by unpaired Wilcoxon test: *p<0.05, **p < 0.01, ***p < 0.001, ****p< 0.0001. EM = effector memory, CM = central memory, EMRA = effector memory cells expressing CD45RA

To characterize B-cell activation states in DLBCL-CAR-T-19 vaccine responders, we analyzed B-cells with 14-marker FlowSOM clustering. We observed distinct differences in B-cell subset distribution between DLBCL-CAR-T-19 vaccine responders and non-responders (**Supplemental Figures 2A, B**). Responders showed enrichment in clusters 2 (naive B cells), 4 (circulating pre-class switched memory B cells), and 6 (non-class switched memory B cells) (**Supplemental Figures 2A-E**). In contrast, non-responders were enriched for cluster 3, early plasmablast precursors (**Supplemental Figures 2B-E**). We observed a positive correlation between time from CART-19 infusion and two B cell subsets (naive and pre-class switched memory B cells), when all patients (composite; responders and non-responders) were analyzed (**Supplemental Figures 3A, C, D**). In the responder cohort, pre-class switched memory B cells (cluster 4), significantly correlated with duration post CART-19 therapy prior to vaccination (**Supplemental Figure 3C**). Together, these data suggest that pre- and non-class switched memory B subsets are associated with the ability to coordinate a vaccine response in DLBCL, and that pre-class switched memory B-cells may represent the most important predictive feature of vaccine response.

### CD4 T cell populations and vaccine response

Next, we analyzed non-naïve CD4^+^ T cells given the importance of T follicular helper (Tfh) cells in generating high affinity antibodies. Indeed, nonresponders showed differences compared to responders and healthy adults (**Figure 5A**). These differences were in regions marked by CXCR5, CD38, CCR7 and CD27, which suggest Tfh cells (**Figures 5A, B**). FlowSOM clustering revealed 2 additional clusters with differential participation of cells in DLBCL-CART-19 vaccine responders compared to the non-responders (clusters 3 and 5, **Figures 5B, D**). In comparison to healthy donors and DLBCL-CART-19 vaccine responders, the frequency of cells in cluster 3 was increased in non-responders, expressing markers resembling cells of Th17-like phenotype (**Figures 5B-E**). In contrast, cluster 5 was increased in vaccine responders, expressing markers consistent with activated circulatory Tfh with a central memory phenotype (**Figures 5B-E**). We also assessed the correlation between the frequency of Tfh and time from CART-19 therapy, as well as B cell subsets enriched in responders or non-responders (**Supplemental Figures 3E-I**). We observed statistically significant positive and negative correlation between non-class switched memory B cells (cluster 6) and early plasmablast precursors (cluster 3) with Tfh frequency (**Supplemental Figures 3G, I**), highlighting the contribution of Tfh and early plasmablast precursors to vaccine response and nonresponse, respectively.

**Figure 5:**
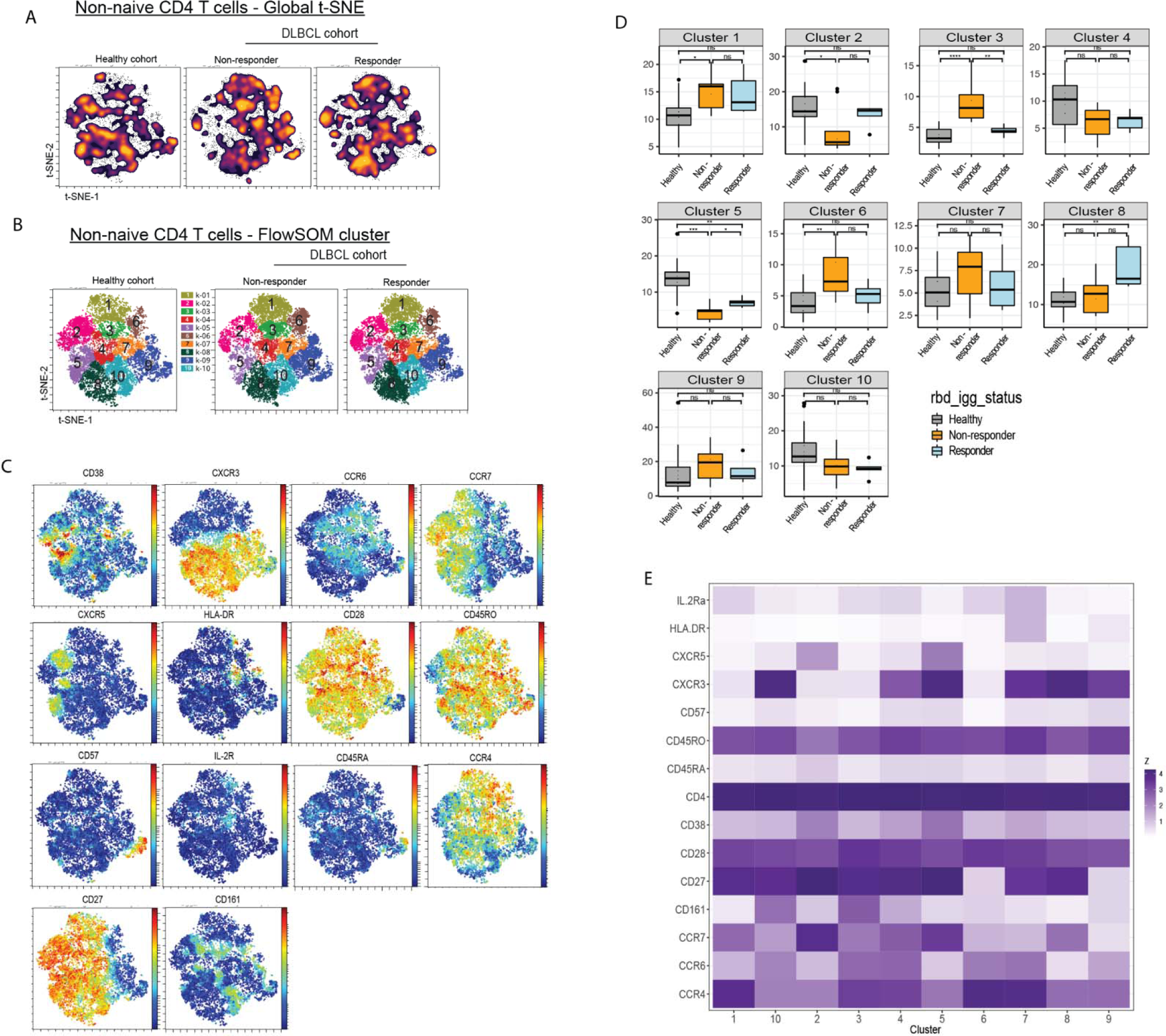
Increased Tfh frequency is associated with vaccine response in DLBCL post CAR T cell therapy. (**A**) Global t-SNE of non-naïve CD4^+^ T cell projection from each indicated cohort. (**B**) FlowSOM cluster of non-naïve CD4^+^ T cells from each indicated cohort. (**C**) Projection of each indicated protein onto the t-SNE map in (a). (**D**) Frequencies of non-naïve CD4^+^ T cells from each cohort in each FlowSOM cluster in (a). Responder (n=5), Non-responder (n=9) and Healthy (n=23). (**E**) MFI of each marker in each FlowSOM cluster (row scaled z-score). Markers expression by cluster 3 (CD161+CXCR3-CCR6-, Th17 cells) and cluster 5 (CCR7+CD38+CXCR3+CXCR5+CD27+CD45RO+, activated circulatory Tfh with a central memory phenotype). Significance was calculated by unpaired Wilcoxon test: *p<0.05, **p < 0.01, ***p < 0.001, ****p< 0.0001

### SARS-CoV-2 mRNA vaccine 3rd dose induced seroconversion in a subset of patients on BTK inhibition

Finally, we assessed serologic anti-RBD IgG responses in a subset of patients with lymphoma/CLL who received a third dose of SARS-CoV-2 mRNA vaccine (**Supplemental Table 4**). In the research cohort, seven patients received a third mRNA vaccine dose and had RBD-specific IgG assessed at a median of 11 days (range: 7-15 days) after vaccination. Only 1 of 7 patients was seropositive before the third dose (**Figure 6A**). Of the remaining 6 patients, 3 (50%) converted from SARS-CoV-2 antibody negative to positive, whereas 3/6 (50%) remained negative for RBD-specific IgG. In the clinical cohort, forty-nine patients received a third mRNA vaccine dose and underwent subsequent clinical assessment of RBD-specific IgG (**Figure 6B**). Prior to the third dose, 17/49 patients (35%) were seropositive. At a median of 5 weeks post third vaccination (range: 0.5-14 weeks), anti-RBD IgG concentration increased. Ten of 32 (31%) seronegative (including equivocal) patients developed detectable RBD-specific IgG. The remaining 22 of 32 (69%) seronegative patients continued to have undetectable or equivocal RBD-specific IgG even after the third vaccine dose.

**Figure 6:**
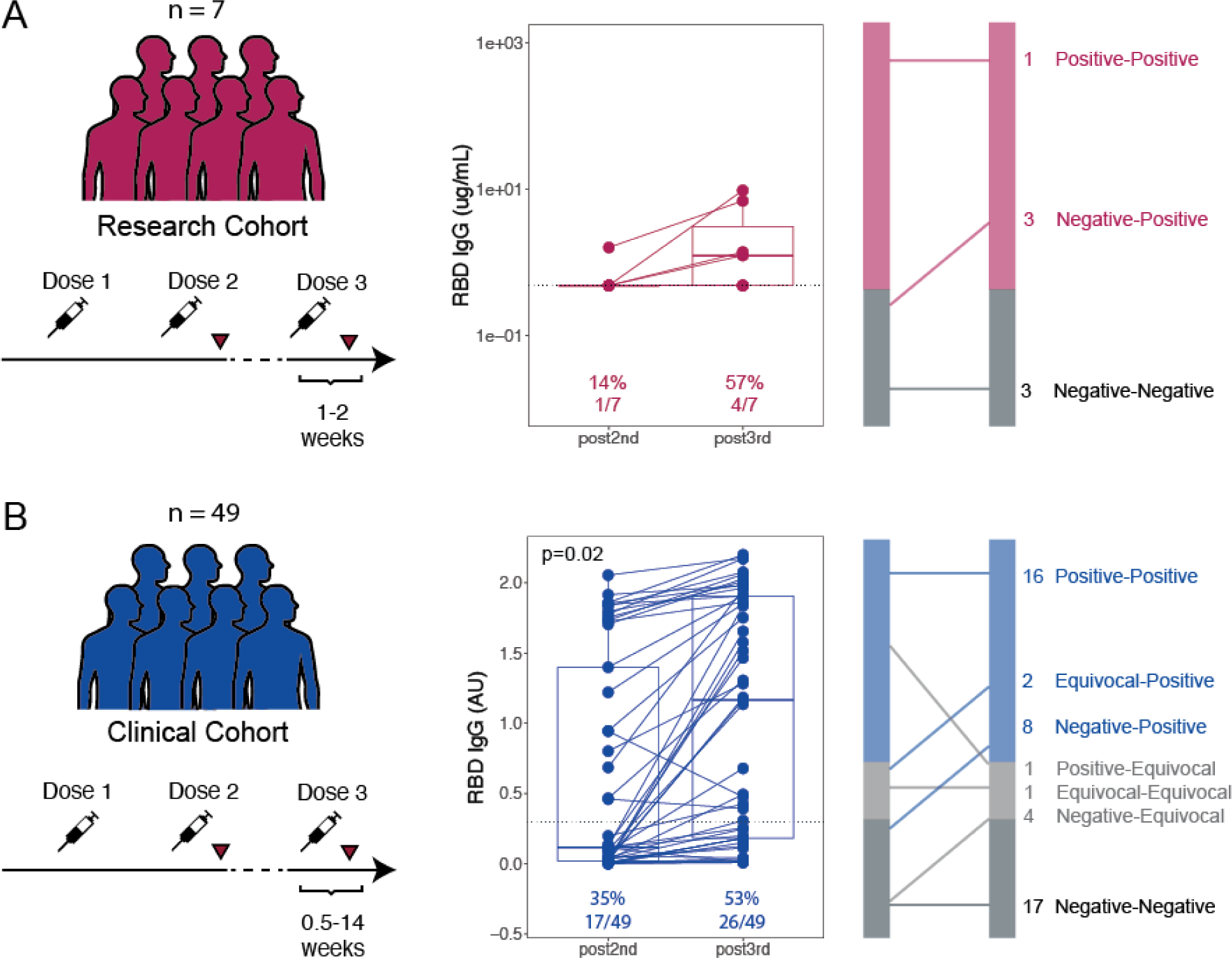
Serologic Response to Third SARS-CoV-2 Vaccination in Patients with Lymphoma/CLL. Paired (**A**) research cohort and (**B**) clinical cohort anti-RBD IgG responses after 2nd and 3rd vaccines doses, with graphical representation of changes at right. P-value derived from Fisher’s exact test.

The disease and treatment status for those who did and did not respond to a 3rd dose of vaccine are listed in **Table 2 and Supplemental Table 6**. Of 56 total patients assessed in the research and clinical cohorts (**Supplemental Table 6**), 54 (96%) maintained the same treatment status (e.g., continued the same therapy or did not receive therapy) between vaccine doses 2 and 3. The majority of patients who converted to positive after receiving a third vaccine dose were not actively receiving treatment (6 of 10, 60%), but 4 (40%) patients were receiving BTKi for iNHL, MCL, or CLL. Of those patients who converted from negative to positive, one patient recovered B cells 15 months after rituximab-chemoimmunotherapy, between second and third vaccination. Only 2 of 22 (9%) patients who remained negative or equivocal after the third dose were not on active therapy, and both had active disease. The only patient who demonstrated a decrease in RBD-specific IgG (titers decreased from positive to equivocal) initiated anti-CD20 therapy between vaccine doses 2 and 3. In summary, across both cohorts, a third dose of vaccine seroconverted 34% (13/38) of patients who were seronegative or equivocal after vaccine dose 2. Patients who derived this benefit were those who were untreated, had completed therapy, or were receiving BTK inhibitors.

**Table 2.**
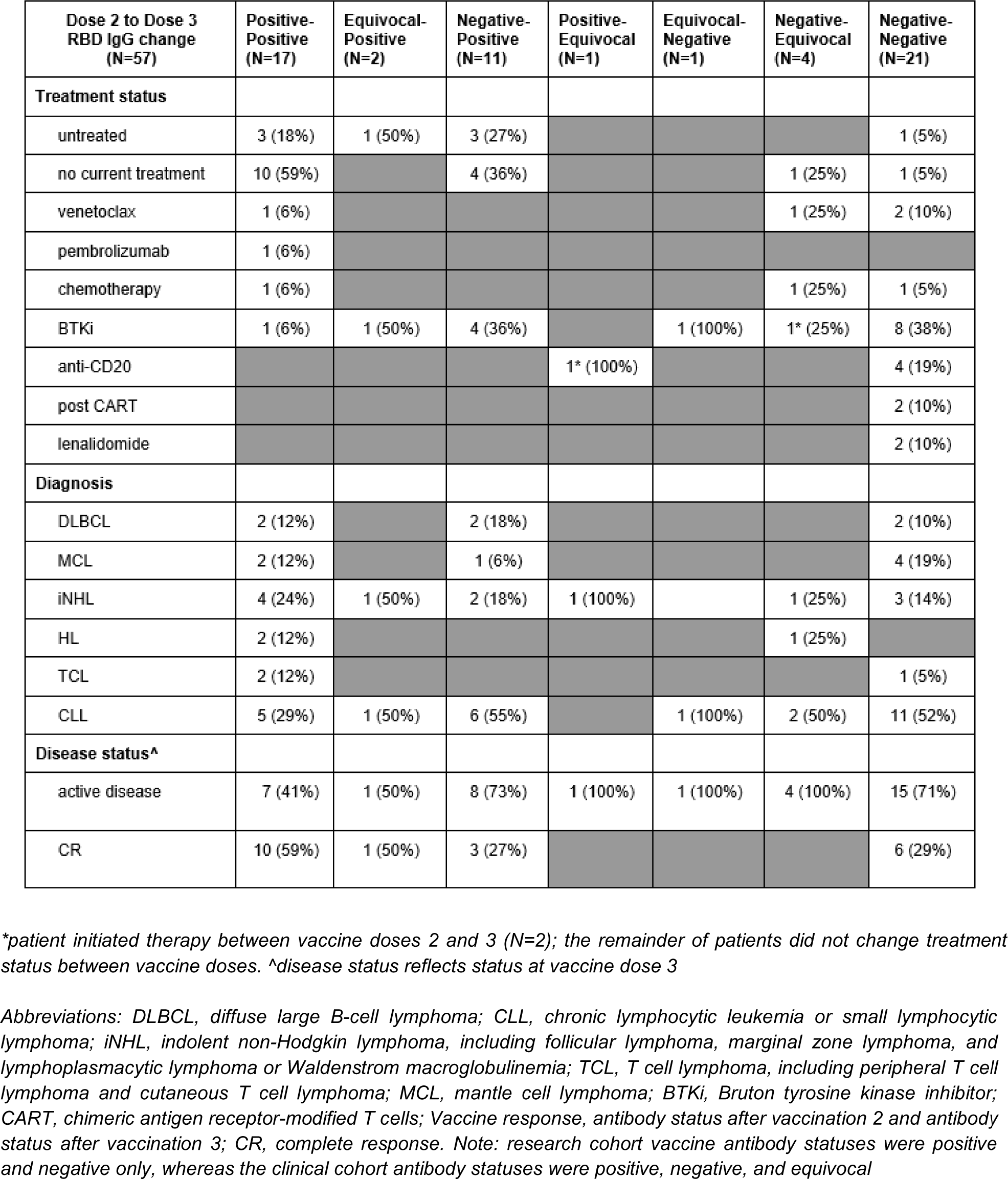
Change in RBD IgG antibody status based on baseline characteristics.

## DISCUSSION

The generation of protective immunity requires a responsive immune system. Studies of vaccine responsiveness in lymphoma prior to 2021 were limited to measuring vaccine responsiveness to recall antigens and therefore studied processes that could draw from memory responses. Here, we leveraged SARS-CoV-2 mRNA vaccination to dissect the clinical and immunologic predictors of *de novo* serologic responsiveness in lymphoma/CLL. We showed serologic response was less frequent during active treatment but that treatment status alone was insufficient to predict responsiveness. When all lymphoma and treatment types were analyzed in composite, total serum levels of IgA and IgM were also weakly correlated with vaccine response. Finally, cellular predictors of vaccine response performed within a uniform population of individuals with DLBCL-CART-19 indicate that, in addition to the frequencies of B cells, the frequency of CXCR5+ T follicular helper-like cells was associated with serologic vaccine responsiveness post-CART-19 therapy.

The presence of active therapy was associated with diminished vaccine response, but non-response also occurred in the absence of active therapy, and even in individuals who had never received treatment, suggesting an adverse impact of intrinsic immunosuppression in lymphoma/CLL. Our data further suggest that the timing of vaccination relative to therapy is an important factor in successful response to vaccination. Consistent with other studies, the effects of anti-CD20 antibodies and chemotherapy were reduced with time, with gradual increases in anti-RBD-specific antibodies as the weeks from anti-CD20 treatment increased ^26,34,35^. Untreated patients may benefit from vaccination at diagnosis or as far in advance of treatment as clinical care allows. However, the time required to initiate and sustain adequate B cell selection and plasma cell formation before initiation of B-cell targeting therapies has not been defined and would likely require several weeks. For patients who received CART-19 therapy, the window of non-response was much longer than the three months post CART-19 currently recommended by published consensus guidelines; instead, vaccine responsiveness may take 1-2 years after CART-19 infusion^36^. Conversely, therapies such as PD-1 blockade had no discernible effect on RBD-specific IgG. The variability in vaccine responsiveness during therapy, observation, and across therapeutic classes suggests that other estimates of vaccine readiness are needed.

Our studies of immunologic predictors of vaccine responsiveness across all groups yielded both cellular and serologic correlates. First, the ability for B cells to class switch and generate IgA and IgM-secreting plasma cells and therefore normal levels of total serum IgA and IgM performed slightly better than absolute T and B cells counts as a correlate of vaccine response. Still, these measures may perform better in some therapy subtypes than others, and the study of clinical immunologic predictors were limited as the labs were not uniformly ordered for clinical care. Unlike clinical immunologic measures, the research cohort more uniformly captured exploratory measures of immune health using CyTOF. When the largest cohort of uniform diagnosis and treatment was studied (DLBCL-CART-19), low or absent B cell frequency was the strongest predictor of vaccine non-response, a finding that would likely be available in clinical B cell counts as well. This observation is consistent with recently published data^37,38^. Beyond B cells, the loss of Tfh in blood was also associated with vaccine non-response. Tfh in blood are recent emigres from lymphoid tissues and may also reflect the end product of productive T and B cell interactions ^39,40^; the presence of circulating Tfh may therefore serve as an additional measure of healthy T and B cell interactions. Of note, the CyTOF studies were performed using only 300ul of whole blood, whereas deep immunophenotyping typically requires cells from 5-10 times that blood volume to fully sample rare populations. Therefore, a more detailed understanding of lymphocytes subsets predictive of vaccine response, particularly in lymphoma subtypes and other conditions where therapies do not primarily deplete B cells, will require study of peripheral blood mononuclear cells in greater depth.

In summary, our study suggests that optimal timing of vaccination in patients with lymphoma may be impacted by lymphoma subtype, treatment received, and time from last therapy. While several immunological measures were correlated with vaccine response, there remains substantial need to develop clinical predictors of immune health and vaccine readiness in all patients. Moving forward, uniform cohorts and deeper analyses of cell types and states are needed to further define the immunologic features that, alone or in combination, predict immunologic vaccine readiness. In turn, these in-depth studies may inform alternative strategies to induce more robust vaccine responses in patients with lymphoma/CLL and achieve actionable measures of immunologic function in settings of immunocompromise.

## DECLARATIONS

### Ethics approval and consent to participate

This research was conducted under an approved protocol by the University of Pennsylvania IRB and in accordance with the Declaration of Helsinki. All patients for the research cohort provided written informed consent. A HIPAA waiver was granted for the clinical cohort.

### Consent for publication

All authors had access to the original data and approved this manuscript for publication.

### Data availability

Requests for data should be directed to the corresponding author. All requests for raw and analyzed pre-clinical data and materials are promptly reviewed by the University of Pennsylvania to determine if they are subject to intellectual property or confidentiality obligations. Patient-related data not included in the paper were generated as part of clinical trials and may be subject to patient confidentiality. Any data and materials that can be shared will be released via a material transfer agreement.

### Author contributions

E.A.C., K.M.W., E.J.W., L.A.V., M.R., S.J.S. designed the research

E.A.C., A.R.G., E.J.W., S.J.S., L.A.V., M.R., S.J.S. oversaw the research

E.A.C., E.R.C., J.F., A.G. collected the data

M.E.W., C.M.M., M.A., N.T., S.G., E.M.D performed the research serology experiments

E.L.P. oversaw the optimization and validation of the clinical lab RBD assays

J.G. and A.G. performed the IgG RBD clinical lab assays

S.E.H. oversaw the optimization and validation of the research RBD assays

K.L. performed the neutralization experiments

P.B. oversaw the optimization and validation of the neutralization assays

E.A.C., K.G.K., G.N.G., S.C.K., J.N., A.L.G., L.A.V. analyzed the data

E.A.C., J.S., S.K.B, E.B.N., R.K.L., S.D.N., J.N.G., D.J.L, S.J.S. managed patients

A.P., A.H., S.K., H.S., S.H., J.T.H, R.P. processed peripheral blood samples

S.A., K.D., J.C.W., A.P., A.H., S.K., H.S., S.H., J.T.H managed the research sample database

E.A.C., K.G.K, M.R., L.A.V. wrote the manuscript

L.A.V., K.G.K., G.N.G, S.C.K., J.N. created figures All authors reviewed and edited the manuscript.

## Acknowledgments

The authors thank Peter and Martha (Muff) Morse whose philanthropy supported this research. The authors also thank all patients and healthy participants, their families and surrogates, as well as the clinical research team and medical personnel who managed patient care.

## ABBREVIATIONS

BTKi: Bruton tyrosine kinase inhibitors
CAR T cells: Anti-CD19 chimeric antigen receptor-modified T Cells
CLL: chronic lymphocytic leukemia/small lymphocytic lymphoma
DLBCL: diffuse large B cell lymphoma
FRNT50: focus reduction neutralization titer 50%
HL: Hodgkin lymphoma
IgG: immunoglobulin G
IgA: immunoglobulin A
IgM: immunoglobulin M
iNHL: indolent non-Hodgkin lymphoma
MCL: mantle cell lymphoma
NHL/CLL: non-Hodgkin lymphomas and chronic lymphocytic leukemia/small lymphocytic lymphoma
PD1: programmed cell death protein 1
RBD: receptor-binding domain
SARS-CoV-2: severe acute respiratory syndrome coronavirus 2
TCL: T cell lymphoma

