## Supplemental Material for "Immunologic predictors of vaccine responsiveness in patients with lymphoma and CLL"

**Supplemental Material for “Title: Immunologic predictors of vaccine responsiveness in patients with lymphoma and CLL”**

**Authors:**

Elise A. Chong, MD^#^*^1,2^, Kingsley Gideon Kumashie*^3^, Emeline R. Chong, BA^1^, Joseph Fabrizio, BS^1^, Aditi Gupta, MD^2^, Jakub Svoboda, MD^1,2^, Stefan K. Barta, MD^1,2^, Kristy M. Walsh, BS^1^, Ellen B. Napier, CRNP^1^, Rachel K. Lundberg, PA-C^1^, Sunita D. Nasta, MD^1,2^, James N. Gerson, MD^1,2^, Daniel J. Landsburg, MD^1,2^, Joyce Gonzalez, BS^4^, Andrew Gaano, BS^4^, Madison E. Weirick, BS^5^, Christopher M. McAllister, BS^5^, Moses Awofolaju, BA^5^, Gavin N. John^3^, Shane C. Kammerman^3^, Josef Novaceck^3^, Raymone Pajarillo, MS^6^, Kendall A. Lundgreen, PhD^5^, Nicole Tanenbaum, BA^5^, Sigrid Gouma, PhD^5^, Elizabeth M. Drapeau, PhD^5^, Sharon Adamski, MS^7,8^, Kurt D’Andrea, BS^7,8^, Ajinkya Pattekar, MD^6,8^, Amanda Hicks, BS^7,8^, Scott Korte, BS^7,8^, Harsh Sharma, MS^7,8^, Sarah Herring, BS^7,8^, Justine C. Williams, BS^7,8^, Jacob T. Hamilton, MS^7,8^, Paul Bates, PhD^5^, Scott E. Hensley, PhD^5^, Eline T. Luning Prak, MD, PhD^4,8^, Allison R. Greenplate, PhD^7,8^, E. John Wherry, PhD^7,8,9^, Stephen J. Schuster, MD* ^1,2^, Marco Ruella, MD*^1,2,6,7^ and Laura A. Vella, MD, PhD* ^3,8^

**SUPPLEMENTAL FIGURES**

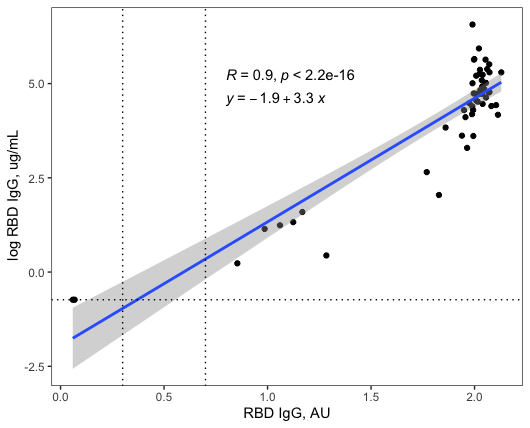

**Supplemental Figure 1 | Comparison of clinical and research RBD results for healthy donors.** Serum samples from healthy donors were divided and both clinical and research RBD assays were performed. P-value calculated using Pearson correlation test.

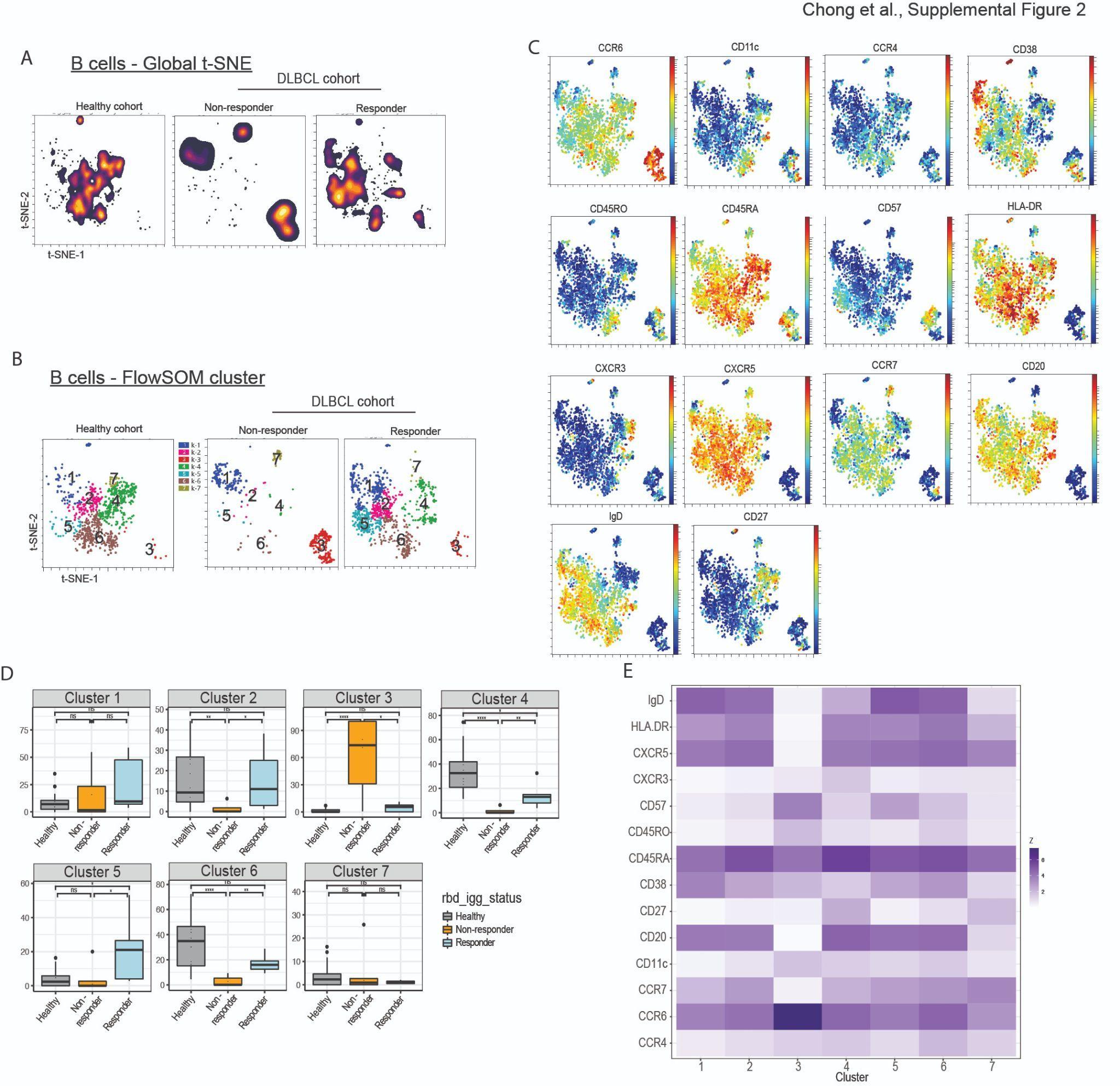

**Supplemental Figure 2: Naive and activated B cells are associated with SARS-CoV-2 mRNA vaccine response in DLBCL post CART-19 therapy**

(**A**) Global t-SNE projection of B cells for each cohort. (**B**) FlowSOM cluster of B cells from each indicated cohort. (**C**) Projection of each indicated protein onto t-SNE map. (**D**) Frequency of B cells from each cohort in each indicated FlowSOM cluster. (**E**) MFI of each marker in each FlowSOM cluster (row scaled z-score). Markers for clusters with significant differences: clusters 2 (IgD^+^CD27^-^), cluster 4 (CD27^+^CD38^dim^IgD^low/-)^, cluster 6 (CD27^dim^CD38^+^IgD^+^) and cluster 3 (CCR6^high^CD20^-^CD27^lo/-^IgD^lo/-^). Significance was calculated using unpaired Wilcoxon test: *p<0.05, **p < 0.01, ****p< 0.0001

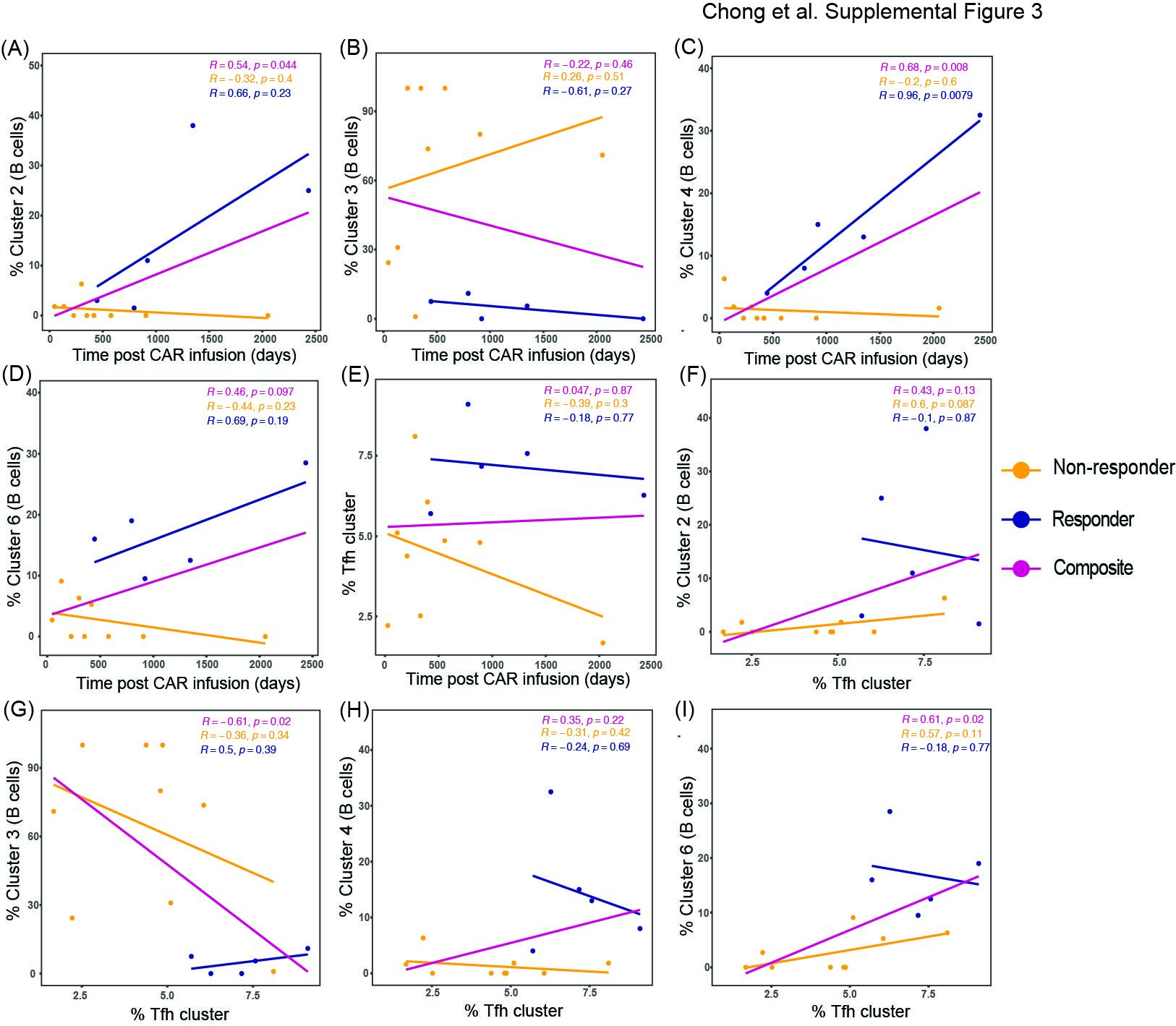

**Supplemental Figure 3: Comparison of the duration of CAR T therapy prior to vaccination, Tfh cells and B cell subsets**

(**A - E**) Correlation analyses of vaccination time point post CAR T cell therapy to B cell subset and Tfh cells. (**F- I**) Comparison of Tfh cell frequencies to B cell subsets enriched in responders or non-responders. P-value calculated using Pearson correlation test.

**Supplemental Table 1| Markers included in defining t-SNE map for the mononuclear cells (lymphocytes and monocytes), B cells, non-naive CD4+ T cells**

| **Immune cell subset** | **Markers** |
| --- | --- |
| Mononuclear cells (Lymphocytes and monocytes) | CCR6, IL-7R (CD127), CCR4, CD14, IgD, CCR7, CD28, CXCR5, CXCR3, CD57, CD27, HLA-DR, CD20, CD161, CD45RO, CD11c, CD294, CD56, CD45RA, CD38, CD8, CD4, CD19, CD14, and IL-3R |
| B cells | IgD, HLA-DR, CD11c, CXCR5, CXCR3, CD57, CD45RO, CD45RA, CD38, CD27, CD20, CCR7, CCR6 and CCR4 |
| Non-naive CD4+ T cells | IL-2R (CD25), HLA-DR, CXCR5, CXCR3, CD57, CD45RO, CD45RA, CD38, CD28, CD27, CD161, CD4, CCR7, CCR6 and CCR4 |

**Supplemental Table 2 | Neutralizing Antibodies: Patients’ Characteristics**

|  | **Research Cohort (N = 14)** | **Healthy Cohort**  **(N=9)** | **Tests for Difference** |
| --- | --- | --- | --- |
| **Age, median (range)** | 66 (45-78) | 46 (41-63) | p = 0.002 |
| **Sex, female, n (%)** | 2 (14) | 6 (67) | p = 0.028 |
| **Vaccine type**  Pfizer  Moderna | 6 (43)  8 (57) | 9 (100)  0 (0) | p = 0.007 |
| **Diagnosis**  DLBCL  CLL  FL  MZL | 3 (21)  5 (36)  3 (21)  3 (21) | -  -  - | -  -  - |
| **Disease status**  active disease  CR | 9 (64)  5 (36) | -  - | -  - |
| **Therapy**  untreated  no current treatment  BTKi  post CART | 8 (57)  2 (14)  2 (14)  2 (14) | -  -  -  - | -  -  -  - |

*Abbreviations: MZL, marginal zone lymphoma; CLL, chronic lymphocytic leukemia; FL, follicular lymphoma; DLBCL, diffuse large B-cell lymphoma; BTKi, Bruton tyrosine kinase inhibitor; CAR T-cells, anti-CD19 chimeric antigen receptor-modified T-cells; CR, complete response.*

**Supplemental Table 3 | CART-19 treated DLBCL patient cohort for CyTOF analysis**

| **Age** | 70s | 60s | 70s | 70s | 50s | 50s | 60s | 60s | 70s | 40s | 40s | 60s | 60s | 60s |
| --- | --- | --- | --- | --- | --- | --- | --- | --- | --- | --- | --- | --- | --- | --- |
| **Disease status** | CR | AD | CR | CR | CR | CR | CR | CR | CR | CR | CR | CR | CR | CR |
| **Vaccine dose (VD) 1 date** | 2/21 | 3/21 | 1/21 | 2/21 | 2/21 | 1/21 | 4/21 | 1/21 | 3/21 | 4/21 | 2/21 | 3/21 | 3/21 | 4/21 |
| **VD2 (rel. to VD1; days)** | +28 | +21 | +28 | +21 | +20 | +28 | +29 | +24 | +28 | +18 | +28 | +28 | +21 | +21 |
| **Blood sample post VD2 (days)** | 31 | 26 | 29 | 46 | 47 | 82 | 10 | 83 | 50 | 40 | 66 | 72 | 124 | 73 |
| **RBD-IgG status** | -ve | -ve | -ve | +ve | -ve | -ve | -ve | +ve | +ve | +ve | -ve | +ve | -ve | -ve |
| **CART19 infusion days prior to VD1** | -134 | -577 | -351 | -921 | -2054 | -417 | -906 | -1348 | -2436 | -795 | -46 | -446 | -300 | -226 |
| **ALC** | 0.2 | 0.7 | 0.6 | 0.8 | 1.79 | 1.23 | 0.6 | 2.3 | 1.4 | 1.1 | 0.7 | 0.9 | 1.2 | 0.2 |
| **CD3 count** | 232 | 732 | 503 | NA | NA | 470 | NA | NA | NA | NA | 76 | 869 | NA | NA |
| **CD4 count** | 149 | 214 | 345 | NA | 742 | 305 | NA | NA | NA | NA | 53 | 544 | 602 | NA |
| **CD8 count** | 93 | 509 | 159 | NA | 660 | 167 | NA | NA | NA | NA | 21 | 298 | 375 | NA |
| **CD19 count** | 9 | 9 | 9 | NA | 1 | NA | 0 | NA | NA | NA | 9 | 19 | 21 | NA |
| **IgG (ug/ml)** | NA | 809 | 635 | NA | NA | 843 | 160 | NA | NA | 309 | 460 | 388 | 509 | NA |
| **IgA (ug/ml)** | 4 | 46 | 76 | NA | 11 | 89 | 9 | NA | NA | 40 | 99 | 44 | 6 | 18 |
| **IgM (ug/ml)** | 9 | 9 | 19 | NA | 11 | 15 | 9 | NA | NA | 19 | 19 | 9 | 9 | 9 |
| **Rx number** | 3 | 4 | 2 | 4 | 7 | 3 | 3 | 4 | 5 | 4 | 2 | 3 | 6 | 7 |

Abbreviation: DLBCL, diffuse large B cell lymphoma; CR, complete remission; CAR, chimeric antigen receptor; RBD, receptor binding domain; Rx, treatment; ALC, absolute lymphocyte count; -ve, negative; +ve, positive; AD, active disease; NA, not available; Rx, treatme

**Supplemental Table 4 | Third Vaccine Dose: Patients’ Characteristics**

|  | **All Patients (N=57)** | **Clinical Cohort (N=49)** | **Research Cohort (N=8)** |
| --- | --- | --- | --- |
| **Diagnosis**  DLBCL  CLL  iNHL  TCL  HL  MCL | 6 (11)  26 (46)  12 (21)  3 (5)  3 (5)  7 (12) | 4 (8)  21 (43)  12 (24)  2 (4)  3 (6)  7 (14) | 2 (25)  5 (63)  -  1 (13)  -  - |
| **Disease status**  active disease  CR | 38 (67)  19 (33) | 32 (65)  17 (35) | 6 (75)  2 (25) |
| **Therapy**  untreated  no current treatment  BTKi  chemotherapy  post CART  rituximab  venetoclax  lenalidomide  anti-PD-1  change in therapy between vax 2 - vax 3 | 8 (14)  16 (28)  15 (26)  3 (5)  2 (4)  4 (7)  4 (7)  2 (4)  1 (2)  2 (4) | 6 (12)  15 (30)  12 (24)  2 (4)  1 (2)  4 (8)  4 (8)  2 (4)  1 (2)  2 (4) | 2 (25)  1 (13)  3 (38)  1 (13)  1 (13)  -  -  -  -  0 (0) |
| **Vaccine response**  **(vax 2 ab - vax 3 ab)** |  |  |  |
| positive-positive | 17 (30) | 16 (33) | 1 (13) |
| negative-positive | 11 (19) | 8 (16) | 3 (38) |
| negative-negative | 21 (37) | 17 (35) | 4 (50) |
| equivocal-positive | 2 (4) | 2 (4) | - |
| negative-equivocal | 4 (7) | 4 (8) | - |
| equivocal-equivocal | 1 (2) | 1 (2) | - |
| positive -equivocal | 1 (2) | 1 (2) | - |

*Abbreviations: DLBCL, diffuse large B-cell lymphoma; CLL, chronic lymphocytic leukemia or small lymphocytic lymphoma; iNHL, indolent non-Hodgkin lymphoma, including follicular lymphoma, marginal zone lymphoma, and lymphoplasmacytic lymphoma or Waldenstrom macroglobulinemia; TCL, T cell lymphoma, including peripheral T cell lymphoma and cutaneous T cell lymphoma; MCL, mantle cell lymphoma; BTKi, Bruton tyrosine kinase inhibitor; CART, chimeric antigen receptor-modified T cells; anti-PD-1, pembrolizumab; Vaccine response, antibody status after vaccination 2 and antibody status after vaccination 3; CR, complete response.*

*Note: research cohort vaccine antibody statuses were positive and negative only, whereas the clinical cohort antibody statuses were positive, negative, and equivocal*

**Supplemental Table 5 | Change in RBD IgG antibody status based on baseline characteristics.**

| **Dose 2 to Dose 3**  **RBD IgG change**  **(N=57)** | **Positive-**  **Positive (N=17)** | **Equivocal-**  **Positive**  **(N=2)** | **Negative-**  **Positive**  **(N=11)** | **Positive-**  **Equivocal (N=1)** | **Equivocal-Negative (N=1)** | **Negative-Equivocal (N=4)** | **Negative-**  **Negative (N=21)** |
| --- | --- | --- | --- | --- | --- | --- | --- |
| **Treatment status** |  |  |  |  |  |  |  |
| untreated | 3 (18%) | 1 (50%) | 3 (27%) |  |  |  | 1 (5%) |
| no current treatment | 10 (59%) |  | 4 (36%) |  |  | 1 (25%) | 1 (5%) |
| venetoclax | 1 (6%) |  |  |  |  | 1 (25%) | 2 (10%) |
| pembrolizumab | 1 (6%) |  |  |  |  |  |  |
| chemotherapy | 1 (6%) |  |  |  |  | 1 (25%) | 1 (5%) |
| BTKi | 1 (6%) | 1 (50%) | 4 (36%) |  | 1 (100%) | 1* (25%) | 8 (38%) |
| anti-CD20 |  |  |  | 1* (100%) |  |  | 4 (19%) |
| post CART |  |  |  |  |  |  | 2 (10%) |
| lenalidomide |  |  |  |  |  |  | 2 (10%) |
| **Diagnosis** |  |  |  |  |  |  |  |
| DLBCL | 2 (12%) |  | 2 (18%) |  |  |  | 2 (10%) |
| MCL | 2 (12%) |  | 1 (6%) |  |  |  | 4 (19%) |
| iNHL | 4 (24%) | 1 (50%) | 2 (18%) | 1 (100%) |  | 1 (25%) | 3 (14%) |
| HL | 2 (12%) |  |  |  |  | 1 (25%) |  |
| TCL | 2 (12%) |  |  |  |  |  | 1 (5%) |
| CLL | 5 (29%) | 1 (50%) | 6 (55%) |  | 1 (100%) | 2 (50%) | 11 (52%) |
| **Disease status^** |  |  |  |  |  |  |  |
| active disease | 7 (41%) | 1 (50%) | 8 (73%) | 1 (100%) | 1 (100%) | 4 (100%) | 15 (71%) |
| CR | 10 (59%) | 1 (50%) | 3 (27%) |  |  |  | 6 (29%) |

*patient initiated therapy between vaccine doses 2 and 3 (N=2); the remainder of patients did not change treatment status between vaccine doses. ^disease status reflects status at vaccine dose 3

*Abbreviations: DLBCL, diffuse large B-cell lymphoma; CLL, chronic lymphocytic leukemia or small lymphocytic lymphoma; iNHL, indolent non-Hodgkin lymphoma, including follicular lymphoma, marginal zone lymphoma, and lymphoplasmacytic lymphoma or Waldenstrom macroglobulinemia; TCL, T cell lymphoma, including peripheral T cell lymphoma and cutaneous T cell lymphoma; MCL, mantle cell lymphoma; BTKi, Bruton tyrosine kinase inhibitor; CART, chimeric antigen receptor-modified T cells; Vaccine response, antibody status after vaccination 2 and antibody status after vaccination 3; CR, complete response.note: research cohort vaccine antibody statuses were positive and negative only, whereas the clinical cohort antibody statuses were positive, negative, and equivocal*

**Supplemental Table 6 | List of all patients who received vaccine dose 3.**

| **Cohort** | **Dose 2 to**  **Dose 3 Result** | **Treatment** | **Group** | **Disease status** |
| --- | --- | --- | --- | --- |
| Clinical | Positive-Positive | no current treatment | DLBCL | CR |
| Clinical | Positive-Positive | no current treatment | DLBCL | CR |
| Clinical | Positive-Positive | no current treatment | MCL | CR |
| Clinical | Positive-Positive | no current treatment | MCL | CR |
| Clinical | Positive-Positive | no current treatment | iNHL | CR |
| Clinical | Positive-Positive | no current treatment | iNHL | CR |
| Clinical | Positive-Positive | no current treatment | iNHL | CR |
| Clinical | Positive-Positive | no current treatment | iNHL | active disease |
| Clinical | Positive-Positive | no current treatment | HL | CR |
| Clinical | Positive-Positive | no current treatment | TCL | CR |
| Clinical | Positive-Positive | untreated | CLL | active disease |
| Clinical | Positive-Positive | untreated | CLL | active disease |
| Clinical | Positive-Positive | untreated | CLL | active disease |
| Clinical | Positive-Positive | venetoclax | CLL | active disease |
| Clinical | Positive-Positive | pembrolizumab | HL | CR |
| Clinical | Positive-Positive | chemotherapy | TCL | active disease |
| Clinical | Equivocal-Positive | untreated | CLL | active disease |
| Clinical | Equivocal-Positive | BTKi | iNHL | active disease |
| Clinical | Negative-Positive | no current treatment | DLBCL | CR |
| Clinical | Negative-Positive | no current treatment | CLL | active disease |
| Clinical | Negative-Positive | no current treatment | CLL | CR |
| Clinical | Negative-Positive | untreated | iNHL | active disease |
| Clinical | Negative-Positive | untreated | CLL | active disease |
| Clinical | Negative-Positive | BTKi | MCL | active disease |
| Clinical | Negative-Positive | BTKi | CLL | active disease |
| Clinical | Negative-Positive | BTKi | iNHL | active disease |
| Clinical | Positive-Equivocal | new therapy - rituximab started | iNHL | active disease |
| Clinical | Equivocal-Equivocal | BTKi | CLL | active disease |
| Clinical | Negative-Equivocal | chemo | HL | active disease |
| Clinical | Negative-Equivocal | venetoclax | CLL | active disease |
| Clinical | Negative-Equivocal | new therapy -  BTKi started | CLL | active disease |
| Clinical | Negative-Equivocal | no current treatment | iNHL | active disease |
| Clinical | Negative-Negative | no current treatment | CLL | active disease |
| Clinical | Negative-Negative | rituximab | MCL | CR |
| Clinical | Negative-Negative | rituximab | iNHL | active disease |
| Clinical | Negative-Negative | rituximab | MCL | CR |
| Clinical | Negative-Negative | rituximab | MCL | CR |
| Clinical | Negative-Negative | BTKi | CLL | active disease |
| Clinical | Negative-Negative | BTKi | CLL | active disease |
| Clinical | Negative-Negative | BTKi | CLL | active disease |
| Clinical | Negative-Negative | BTKi | CLL | active disease |
| Clinical | Negative-Negative | BTKi | CLL | active disease |
| Clinical | Negative-Negative | BTKi | CLL | active disease |
| Clinical | Negative-Negative | BTKi | iNHL | active disease |
| Clinical | Negative-Negative | venetoclax | CLL | active disease |
| Clinical | Negative-Negative | venetoclax | CLL | active disease |
| Clinical | Negative-Negative | lenalidomide | MCL | CR |
| Clinical | Negative-Negative | lenalidomide | DLBCL | active disease |
| Clinical | Negative-Negative | post CART | iNHL | CR |
| Research | Negative-Negative | post CART | DLBCL | CR |
| Research | Negative-Negative | untreated | CLL | active disease |
| Research | Negative-Negative | BTKi | CLL | active disease |
| Research | Negative-Negative | chemo | PTCL | active disease |
| Research | Negative-Positive | BTKi | CLL | active disease |

| Research | Negative-Positive | untreated | CLL | active disease |
| --- | --- | --- | --- | --- |
| Research | Negative-Positive | no current treatment | DLBCL | CR |
| Research | Positive-Positive | BTKi | CLL | active disease |

**SUPPLEMENTAL METHODS**

***Patients and healthy donors***

Prospective Research Cohort: Sample collection spanned February 15, 2021 to September 8, 2021. Patients had a diagnosis of indolent NHL or CLL and were either untreated, receiving BTK inhibitors, receiving or status post rituximab-chemotherapy, or status post CAR T cells. At enrollment, all patients planned to receive or were within 1 month of receipt of SARS-CoV-2 vaccination. Healthy Cohort: Sample collection occurred from December 21, 2020 to March 29, 2021. Retrospective Clinical Cohort: Data were collected between January 1, 2020 and November 21, 2021. Patients were vaccinated against SARS-CoV-2 by July 12, 2021 and had SARS-CoV-2 antibody levels in their electronic medical record. Patients with documented prior SARS-CoV-2 infection or antibody positivity before first vaccination were excluded from the analyses. Written informed consent was obtained for the prospective cohorts, and a waiver of informed consent and HIPAA authorization for the retrospective cohort was obtained. Sample sizes were dictated by patients who met the above criteria, and, in the case of the prospective research cohort, were those patients who consented to research during the time samples were collected.

***Clinical Data Abstraction***

Patient level clinical data were abstracted from the electronic medical record by the investigators. Abstracted variables included diagnosis; treatment history; disease status; vaccination dates and manufacturer; intravenous immunoglobulin (IVIG) use; history of prior COVID-19; anti-receptor-binding domain (RBD) IgG levels; absolute lymphocyte count; absolute CD3, CD4, CD8, and CD19 cell counts; and quantitative IgM, IgG, and IgA levels. Clinical laboratory results were abstracted from the most recent laboratory studies within 6 months prior to vaccination as long as no change in therapy occurred between time of laboratory studies and vaccination. “Low” clinical laboratory values were defined as a value less than the lower limit of normal for the assay. If a patient was receiving IVIG at the time of vaccination, serum IgG values were omitted, but IgG level was classified as “low”. Patients with missing clinical data for a given variable were not included for that particular analysis. Active treatment was defined as vaccination within 5 half-lives of the therapeutic agent, within 3 months of the last chemotherapy dose, within 6 months of the last anti-CD20 antibody dose, or status post CAR T-cells without progressive lymphoma.

***Research and Healthy Cohort Blood Collection and Serology***

Blood was collected into sodium heparin tubes and centrifuged at room temperature to obtain plasma. An Enzyme-Linked Immunosorbent Assay (ELISA) was performed using plates coated with 2ug/mL of the spike protein or the RBD of the spike protein, as described ^29,30^. Plasma samples were diluted 1:50. If the IgG concentration was above the 0.48 ug/mL lower limit of detection (set at 0.48 ug/mL), the sample was re-run in at least a 7-point dilution series for quantitation. A dilution series of the monoclonal SARS-CoV-2 spike specific IgG antibody, IgG CR3022 (specific for the SARS-CoV-2 spike protein) was used as a control across plates and used to convert optical density values into relative antibody concentrations. Plasmids encoding CR3022 were provided by I. Wilson (Scripps)^31^. Assessment of neutralizing antibodies was performed as previously described^28^. SARS-CoV-2 spike proteins analyzed included the alpha (D614G) and delta (B.1.617.2) variants.

***Clinical Cohort Blood Collection and Serology***

Samples were tested for IgG that binds SARS-CoV-2 spike protein RBD in an enzyme-linked immunosorbent assay that was developed in the Clinical Immunology Laboratory of the Hospital of the University of Pennsylvania. This assay used the same RBD antigen as the research assay and was performed with serum at 1:100 dilution on an automated instrument (DYNEX DSX, Dynex Technologies) (**Supplemental Figure 1**).

***Whole Blood CyTOF***

Whole blood (300µl) sampled between 14 days and 4 months after the second dose of vaccination was added to the Maxpar Direct Immune Profiling Assay (MDIPA) kit (Standard BioTools, Cat. # 201334) and mixed. Blood was incubated for 30 minutes in the dark at room temperature. Smart tube Proteomic stabilizer (420µl, Fisher Scientific) was then added to the stained blood and incubated for another 10 minutes in the dark. The stained whole blood along with the stabilizer was then transferred to a cryovial and frozen to -80°C in a controlled rate freezing container. On the day prior to acquisition the cryovials were thawed at 4°C for 30 minutes. Each stained sample was then transferred added to 3ml of 1x Smart tube thaw-lyse buffer and incubated for 10 minutes. Each sample were then centrifuged at 700g * 7 mins, and supernatant aspirated and. T the pellet was then resuspended in 25ml of 1x thaw-lyse buffer forand incubated for 10 minutes incubation. Following another spin, the cells were then resuspended in 125nm Iridium in Maxpar Fix and Perm Buffer (Fluidigm) and incubated overnight. The cells were then washed twice in Maxpar cell staining buffer (Fluidigm) and acquired on the Fluidigm Helios instrument in maxpar cell acquisition solution (Fluidigm).

***High dimensional CyTOF data analysis***

Gated populations of mononuclear cells (lymphocytes and monocytes; CD45+CD66b-), B cells (CD3-CD19+) and non-naive CD4^+^ (CD4^+^ T cells not co-expressing CD27 and CD45RA) T cells were analyzed separately. t-distributed stochastic neighbor embedding (t-SNE) analysis was performed for all cell subsets using the following parameters:using 6,500 mononuclear cells per sample,. For B cells, 1,000 B cells per study cohort and 15,000 non-naive CD8^+^ and CD4^+^ T cells an equal number of cells were subsampled per study cohort. for analysis: 15,000 each for non-naïve CD4^+^ and 1,000 B cells. All t-SNE analyses were performed with the following settings: 1,000 iterations, 250 early exaggeration iter, 5,000 learning rate, 30 perplexity and 0.5 theta. The markers used to perform the analyses in defining t-SNE analysesmaps for each cell subset are listed in **Supplemental** **Table 1**. The resulting t-SNE map was fed into the FlowSOM clustering algorithm in addition to the markers used in defining the t-SNE map for each cell subset and analyzed. On each cell subset, and a new self-organizing map was generated using Elbow meta-clustering on the t-SNE axis. We observed 9 clusters for mononuclear cells, 10 clusters for non-naive CD4^+^ and CD8^+^ T cells, and 7 clusters for B cells.

***Statistical analyses***

All comparisons between two groups of continuous variables were performed using non-parametric testing with Wilcoxon rank sum testing. All comparisons between two categorical variables utilized Fisher exact testing. Comparisons between more than two groups were performed by Kruskal-Wallis testing. Bonferroni correction was utilized for multiple comparisons. All results are represented as the medians unless otherwise indicated. Correlations between SARS-CoV-2-specific antibody types were performed using Pearson correlation coefficients. Correlations between measured clinical immunologic parameters and SARS-CoV-2-specific IgG were performed using Spearman correlation coefficients. Statistical analyses of cluster participation differences between cohorts were performed using the Wilcoxon test. All statistical analyses were performed in R and visualized in RStudio
